# Collaborative and privacy-preserving workflows on a clinical data warehouse: an example developing natural language processing pipelines to detect medical conditions

**DOI:** 10.1101/2023.09.11.23295069

**Authors:** Thomas Petit-Jean, Christel Gérardin, Emmanuelle Berthelot, Gilles Chatellier, Marie Frank, Xavier Tannier, Emmanuelle Kempf, Romain Bey

## Abstract

**Objective:** To develop and validate advanced natural language processing pipelines that detect 18 conditions in clinical notes written in French, among which 16 comorbidities of the Charlson index, while exploring a collaborative and privacy-preserving workflow.

**Materials and methods:** The detection pipelines relied both on rule-based and machine learning algorithms for named entity recognition and entity qualification, respectively. We used a large language model pre-trained on millions of clinical notes along with clinical notes annotated in the context of three cohort studies related to oncology, cardiology and rheumatology, respectively. The overall workflow was conceived to foster collaboration between studies while complying to the privacy constraints of the data warehouse. We estimated the added values of both the advanced technologies and the collaborative setting.

**Results:** The 18 pipelines reached macro-averaged F1-score positive predictive value, sensitivity and specificity of 95.7 (95%CI 94.5 - 96.3), 95.4 (95%CI 94.0 - 96.3), 96.0 (95%CI 94.0 - 96.7) and 99.2 (95%CI 99.0 - 99.4), respectively. F1-scores were superior to those observed using either alternative technologies or non-collaborative settings. The models were shared through a secured registry.

**Conclusions:** We demonstrated that a community of investigators working on a common clinical data warehouse could efficiently and securely collaborate to develop, validate and use sensitive artificial intelligence models. In particular, we provided efficient and robust natural language processing pipelines that detect conditions mentioned in clinical notes.

## 1 INTRODUCTION

Recent scientific breakthroughs demonstrated the high potential of large health databases to generate new medical knowledge but many challenges still remain to structure the emerging research and innovation ecosystems.^[1,2]^ These communities should exploit workflows which judiciously make use of new technologies to leverage data at scale while complying with many constraints that limits data access to protect patients’ privacy. Clinical data warehouses (CDW) are new platforms that enforce these constraints by providing both secured technical environments and dedicated regulatory pathways. CDW’s architectures may themselves benefit from innovations as no single privacy-enhancing technology provides a silver-bullet solution yet.^[3]^ These needs for a workflow’s and architecture’s innovation became even more obvious with the advent of new machine learning models such as large language models or foundation models. Indeed, training these models often requires accessing unminimized datasets, and models are themselves privacy-sensitive as they may leak information related to patients’ records.^[2,4,5]^

These opportunities and challenges can be exemplified by algorithms that automatically detect conditions in clinical notes. Conditions -especially comorbidities-are routinely considered in clinical practice and in epidemiological studies as they identify disease combinations that may require a specific therapeutic approach or are required for a proper population analysis.^[6–8]^ The Charlson Comorbidity Index (CCI) can for instance be computed out of 16 conditions to predict the 1-year mortality of patients admitted in hospitals.^[9]^ Algorithms were consequently proposed to detect those conditions using structured claim data.^[10,11]^ However, this data source being primarily designed for economic purposes, it suffers from missingness issues. Nevertheless, it was shown that information could be efficiently obtained from clinical notes using Natural Language Processing (NLP) algorithms instead, those algorithms relying even more on machine learning (ML) techniques such as language models.^[12–19]^ Developing tools to this end remains challenging, and many difficulties are yet to be overcome for a wide community to benefit from them.^[4,7,19–24]^

First, the optimal NLP technologies are still debated. Although ML approaches have been proven efficient for some tasks, an architecture based on the transposition of expert clinical knowledge in explicit rules still often reaches similar or better performances.^[7,25]^ Hybrid approaches combining rule-based and ML algorithms are a promising field of research in order to benefit from the strengths of both techniques and optimally leverage the expertise of clinicians and computer scientists.^[26]^ Additionally, sharing hybrid models is facilitated as privacy sensitive and non-sensitive parts can be kept distinct. Second, NLP algorithms developed in a study are often difficult to apply to new contexts. Beyond the obvious challenge of transposing algorithms to other languages, each specialty domain may moreover feature specific syntaxes and vocabularies that impact the algorithms’ robustness.^[21,27]^ Consequently, it appears judicious to develop models jointly considering different datasets to enhance their generalizability. Third, it would be relevant to associate various medical expertises to the development of these algorithms but data accesses of investigators are often restricted to the disease-specific cohorts corresponding to their areas of research. Therefore, new privacy-preserving workflows should be implemented to enable large communities to efficiently collaborate.^[5,7,16,28,29]^

To address these entangled technical and organizational issues, we developed, validated and shared hybrid NLP pipelines that detect in clinical notes written in French the 16 conditions of the CCI, extended by the tobacco and alcohol consumption status. Leveraging the hybrid architecture of those pipelines, we adopted a workflow that enabled the association of investigators working on three disease-specific cohorts while complying to the privacy constraints of the clinical data warehouse. We estimated the performance gains provided respectively by the advanced natural language processing technologies and by the collaborative setting.

## 2 MATERIALS AND METHODS

Our study involved clinicians and data from three retrospective observational studies reviewed and approved by the institutional review board of our institution (*IRB00011591*, decisions *CSE18-32, CSE20-55* and *CSE20-93*) and reused a large language model trained on 21 millions pseudonymised clinical notes in the context of a former study (decision *CSE19-20*).^[20]^ French regulation does not require the patient’s written consent for this kind of research, but the patients were informed about this research and those who objected to the secondary use of their data were excluded from the study.^[30]^ Data was pseudonymized by replacing names and places of residence by aliases. We followed the REporting of studies Conducted using Observational Routinely-collected health Data reporting guideline (see checklist in Supplementary Materials).^[31]^

### 2.1 Rheumatology, oncology and cardiology cohorts

Each study’s dataset is constituted by an extraction of the overall database containing routine data of patients cared for in the *Greater Paris University Hospitals* (Assistance Publique-Hôpitaux de Paris, AP-HP, see Supplementary Appendix A for details on cohort selection). To limit privacy exposure, each investigator accessed only her study’s data (see Figure 1).

**Figure 1:**
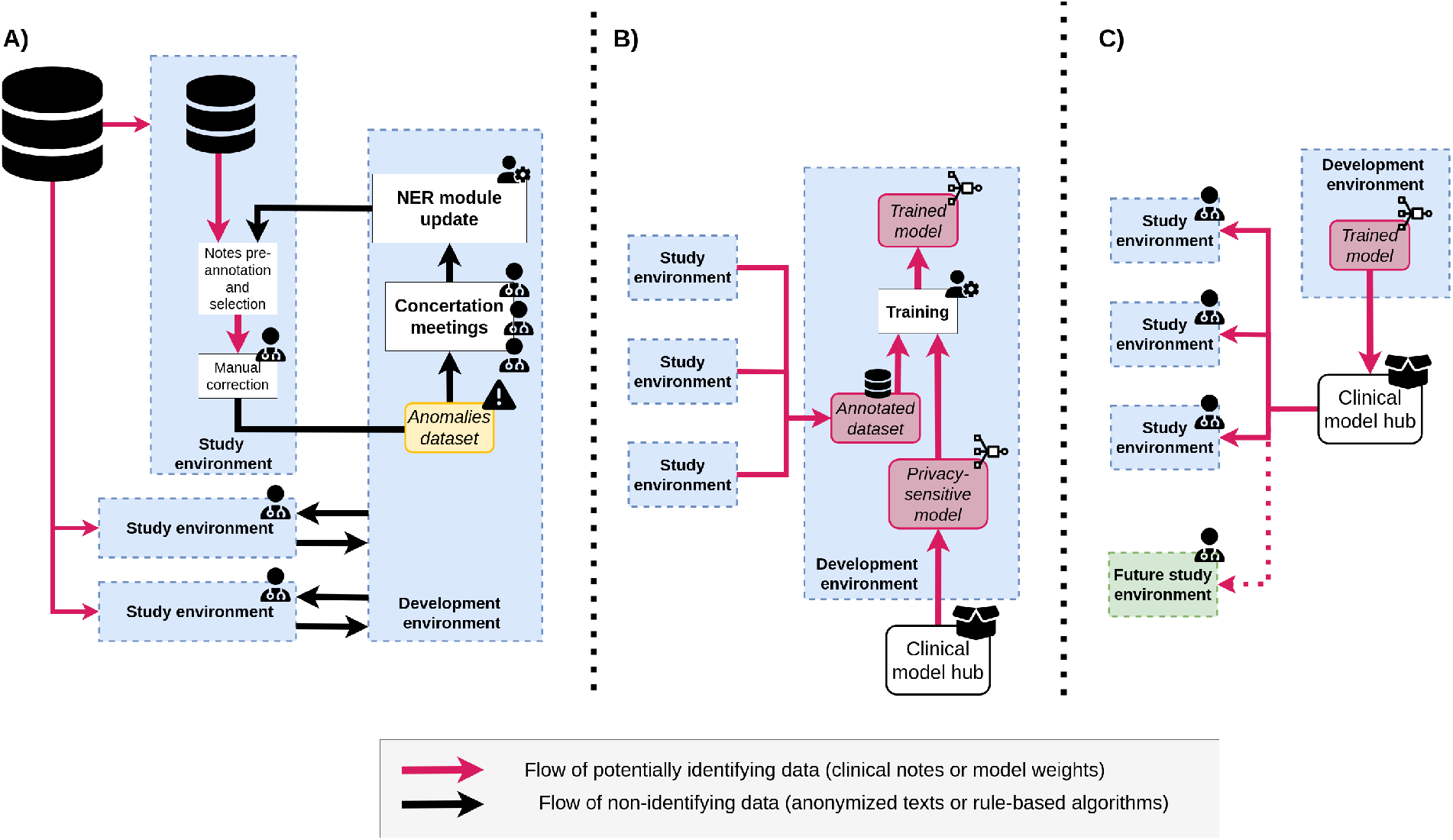
Development, training and serving workflows. **A) Iterative development of the rule-based Named Entity Recognition (NER) algorithm of the Natural Language Processing (NLP) pipeline:** annotation takes place in each study’s secure environment, while development occurs in a separate dedicated environment. **B) Training of the Machine Learning (ML) based qualification algorithm:** the algorithm is initialized by a clinical privacy sensitive model and trained on snippets gathered during A). **C) Validation and serving of the algorithm:** Once trained, the qualification algorithm is pushed to the clinical model hub, from which it can be served to study environments.

We considered demographic data (age at admission, gender), diagnostic claim codes, discharge summaries (for outpatient stays) and consultation reports (for outpatient stays). To avoid data leakage, we followed *Bey et al*. methodology and divided each cohort into a training cohort and a validation cohort (see Supplementary Figure S1).^[32]^

### 2.2 Data sources

AP-HP comprises 38 university hospitals spread across the Paris area (more than 22,000 beds, 1.5 million hospitalizations each year) which use a common electronic health record (EHR) software (ORBIS, Dedalus Healthcare). Data collected in the EHR and in the claim database (French PMSI, *Programme de Médicalisation des Systèmes d’Information*, coded following the ICD-10, the *International Classification of Diseases 10*^*th*^ *revision*) are integrated in the AP-HP CDW on a daily and monthly basis, respectively. The research database follows the *Informatics for Integrating Biology & the Bedside* standard.^[33]^ Data was extracted for this study on April 2^nd^, 2021, July 13^th^, 2021 and October 10^th^, 2021 for the rheumatology, cardiology and oncology cohorts, respectively.

### 2.3 Conditions definitions

We considered the 16 conditions of the CCI. Separately, we also extracted the tobacco and alcohol consumption status. These additions followed both a medical rationale (i.e., the variables were deemed clinically relevant although they would not fit in the original CCI) and an opportunistic rationale (i.e., to make the most of the annotation process). For consistency, they were not considered when computing the CCI. The definitions of these conditions were refined by clinicians (C.G., E.K., E.B.) to optimally balance their relevance for epidemiological research (e.g., avoiding asymptomatic conditions) and the reliability of their detection in clinical notes (e.g., avoiding excessive interpretation of textual mentions). Each condition was defined as a free text, a list of ICD-10 codes inspired by *Sundararajan et al*. and a machine-readable dictionary of synonyms expressed as regular expressions with composition rules (see Supplementary Figure S3).^[11]^ When present, five conditions displayed two possible statuses, namely diabetes (With end-organ damage / Uncomplicated), liver disease (Moderate to severe / Mild), solid tumor (Metastatic / Localized), and tobacco or alcohol consumption (Present / Stopped). An annotation guideline details the annotation methodology (see Supplementary Appendix E).

### 2.4 Pipelines’ architecture

We refer to the main NLP pipeline that we developed as **NLP-ML-CLINICAL**. It consisted of 18 Named Entity Recognition (NER) algorithms that extract mentions of each condition followed by common qualification and aggregation algorithms (see Supplementary Figure S2).

Each **NER algorithm** relied on a curated dictionary of regular expressions for each condition. Additional rules were implemented to deduce, if applicable, the conditions severity from the entity’s context (i.e., a snippet of words around the extraction, the size of which varies between algorithms) and to discard false positives caused by identified mechanisms (e.g., an incorrect acronym). Algorithms were developed within the open source EDS-NLP scientific library and were made available in its 0.8.1 release.^[34]^

The **Qualification algorithm** was a machine-learning module responsible for discarding irrelevant entities. During annotation, entities were labeled as irrelevant if they were either negated (*the patient has no diabetes*), hypothetical (*the patient may have diabetes*) or not related to the patient (*the patient’s father had diabetes*). Generic false positives (e.g., a condition mentioned in a care site name or in a laboratory measure denomination) were also labeled as irrelevant. The algorithm was based on EDS-CamemBERT, a clinical language model that was trained on 21 millions pseudonymised clinical notes from the AP-HP’s CDW.^[20]^ This architecture took a snippet of text containing an entity as input and predicted if the entity should be discarded or not (See Supplementary Appendix B for implementation details).

The **Aggregation algorithm** predicted the conditions at the stay level out of the list of detected and qualified entities in its associated clinical note. For binary predictions, the algorithm predicted the presence of the condition if at least one entity was detected without being discarded. For severity predictions, the algorithm predicted the status of the most severe detected entity.

For comparison, we also developed a set of alternative pipelines. The **NLP-ML-GENERIC** pipeline replaced the EDS-CamemBERT model from **NLP-ML-CLINICAL** with CamemBERT, a publicly available and generic language model.^[35]^ The **NLP-RB** pipeline explored a rule-based approach for the qualification algorithm, implementing three separate algorithms to detect negation, hypothesis and familial relations. Finally, for inpatient stays only, we developed the **CLAIM** pipeline which extracted conditions from claim codes (See Supplementary Appendix B).

### 2.5 Pipelines’ development and training

The NER algorithms and annotations guidelines were jointly initialized by the three expert clinicians (C.G., E.K., E.B.) and the data scientist (T. P.-J.). Two annotation sessions were then organized to improve the pipeline’s performance. At the beginning of each session we used **NLP-RB**, which did not require any training and is not privacy sensitive, to pre-annotate clinical notes. A subset of clinical notes were randomly selected in the training cohorts adopting an upsampling strategy that optimized their relevance (see Supplementary Appendix C). Each clinician read the selected notes relative to her disease-specific cohort and confirmed or infirmed the entity-level predictions while adding entities that would have been falsely omitted by the algorithms. At the end of each session, concertation meetings were organized to discuss clinical and methodological issues, and to improve the conditions definitions and dictionaries. Improvements could include, but were not limited to, adding diseases’ synonyms, performing acronym disambiguation or refining the medical rationale. Only limited, non-identifying anomalies were shared during these meetings (see Figure 1.A).

Once the two annotation sessions were completed, the annotated entities and their contexts were reviewed to fit the consolidated definitions. The three annotated datasets and the pre-trained clinical language model were sent to a common development environment accessed by the data scientist to train the ML qualification algorithm (see Figure 1.B). The clinical language model was imported from a private and secure clinical model hub as it may be considered as personally identifying data.

### 2.6 Pipelines’ validation

The source code and trained parameters of the NLP pipelines were frozen at the beginning of the validation session and the qualification algorithm was made available through the clinical model hub. The validation sub-cohorts were divided into inpatient and outpatient stays. The pipeline was applied to pre-annotate either the last-edited discharge summaries of inpatient stays or the consultation reports of outpatient stays. Each clinician reviewed notes drawn randomly in her disease-specific sub-cohort. After the validation session, two conditions (hemiplegia and AIDS) were found in less than ten clinical notes overall. For those two conditions, we performed when possible an additional validation step by upsampling up to 30 clinical notes on which the condition was extracted by the algorithm. Due to this biased notes selection, we only reported the positive predictive value (PPV) for these conditions. They were excluded from the aggregated metrics.

For every other condition, we reported the F1-score, the PPV, the sensitivity and the specificity. In the case of conditions with two possible statuses, we considered the extraction of each status as a separate binary algorithm and also reported the performance of the binary algorithm consisting of detecting the presence of the condition, whatever its status. Entity-level (i.e., the pipeline minus the aggregation algorithm) and stay-level (i.e., the full pipeline) predictions were both confirmed or invalidated by a chart review of the analyzed notes, thus leading to both an entity-level and a stay-level gold standard dataset. Regarding entity-level performances, we considered a prediction as correct if it overlapped with a gold standard entity.

We moreover assessed the usability of these pipelines to compute the CCI score. Indeed, although its computation is not the objective of the study, it may be realized using the delivered pipelines. We compared the CCI computed using **NLP-ML-CLINICAL** with the same score computed using either the **CLAIM** pipelines or with its value directly mentioned in a subset of ICU notes by the clinicians at the point of care (see Supplementary Appendix D).

### 2.7 Comparison with alternative technologies and collaboration settings

For each condition we compared the performance of **NLP-ML-CLINICAL** with the three alternative pipelines **NLP-ML-GENERIC, NLP-RB** and **CLAIM**. We also evaluated the added value of the collaborative setting on the qualification algorithm by training and validating **NLP-ML-CLINICAL** separately in the three study environments. These performances were compared on the inpatient and outpatient stays of the validation sub-cohorts. Finally, to assess the generalizability of this pipeline, we evaluated its performance when trained and validated on distinct disease-specific cohorts.

## 3 RESULTS

Table 1 shows the number of stays and demographic characteristics of the validation sub-cohorts. Overall, 134 (89%) discharge summaries had at least one valid condition mentioned, on which an average of 10.1 entities were present. Similarly, 112 (75%) consultation reports had at least one valid condition mentioned, with 3.7 entities on average. When also considering discarded entities (e.g., negated or hypothetical), those numbers of discharge summaries and consultation reports increased to 143 and 119, respectively, and the corresponding average number of entities increased to 13.2 and 4.4, respectively (Supplementary Table S4). Regarding discharge summary, routinely reported conditions such as alcohol or tobacco consumption featured the highest increase, from 14 and 35 to 45 and 68, respectively.

**Table 1:** Composition of the validation dataset sampled in the three disease-specific cohorts. For each condition, the number of clinical notes with at least one validated entity is shown along with, for those notes, the mean number of validated entities per note (in brackets). For instance, in the cardiology cohort, 12 discharge summaries mention a myocardial infarction and on average 3.3 validated entities are present within each one of them.

| Stay type | Oncology cohort |  | Cardiology cohort |  | Rheumatology cohort |  | Overall cohort |  |
| --- | --- | --- | --- | --- | --- | --- | --- | --- |
|  | inpatient | outpatient | inpatient | outpatient | inpatient | outpatient | inpatient | outpatient |
| Age at admission | 84.6 (7.3) | 80.9 (6.9) | 61.4 (18.7) | 61.9 (15.2) | 57.5 (19.4) | 53.4 (19.2) | 67.8 (20.0) | 65.4 (18.6) |
| Gender distribution (M-F) | 48 - 52 | 48 - 52 | 60 - 40 | 46 - 54 | 50 - 50 | 36 - 64 | 52 - 47 | 43 - 56 |
| Total annotated records | 50 | 50 | 50 | 50 | 50 | 50 | 50 | 50 |
| Myocardial infarction | 12 (3.3) | 9 (2.1) | 8 (3.0) | 2 (1.0) | 8 (3.1) | 2 (2.0) | 28 (3.2) | 13 (1.9) |
| Congestive heart failure | 34 (6.0) | 21 (2.7) | 11 (4.5) | 1 (4.0) | 13 (4.9) | 2 (1.0) | 58 (5.5) | 24 (2.6) |
| Peripheral vascular disease | 37 (1.8) | 17 (1.9) | 25 (2.2) | 7 (1.3) | 28 (2.5) | 10 (2.2) | 90 (2.1) | 34 (1.9) |
| Cerebrovascular disease | 9 (1.7) | 4 (1.8) | 3 (1.7) | 2 (1.5) | 12 (4.9) | 3 (1.3) | 24 (3.3) | 9 (1.6) |
| Dementia | 13 (1.6) | 2 (2.5) | 3 (1.3) | 0 | 9 (2.8) | 3 (1.3) | 25 (2.0) | 5 (1.8) |
| Chronic pulmonary disease | 18 (3.9) | 14 (1.6) | 5 (3.8) | 4 (1.5) | 10 (4.2) | 7 (1.7) | 33 (4.0) | 25 (1.6) |
| Rheumatologic disease | 1 (1.0) | 3 (1.3) | 3 (2.7) | 1 (2.0) | 13 (4.5) | 12 (1.7) | 17 (4.0) | 16 (1.6) |
| Peptic ulcer disease | 6 (1.3) | 0 | 3 (1.7) | 1 (1.0) | 2 (1.0) | 0 | 11 (1.4) | 1 (1.0) |
| Liver disease | 0 | 1 (3.0) | 2 (4.5) | 1 (2.0) | 7 (5.0) | 2 (2.5) | 9 (4.9) | 4 (2.5) |
| Diabetes | 15 (1.9) | 13 (1.3) | 12 (1.9) | 4 (1.8) | 10 (3.8) | 5 (1.6) | 37 (2.4) | 22 (1.5) |
| Hemiplegia* | 1 (2.0) | 1 (2.0) | 1 + 10 (4.0) | 0 + 13 (-) | 2 + 11 (1.0) | 0 + 12 (-) | 4 + 21 (2.0) | 1 + 25 (2.0) |
| Renal disease | 9 (1.9) | 4 (2.0) | 5 (1.2) | 1 (3.0) | 1 (3.0) | 0 | 15 (1.7) | 5 (2.2) |
| Solid tumor | 7 (2.7) | 6 (2.0) | 21 (3.1) | 21 (2.0) | 7 (1.6) | 6 (1.5) | 35 (2.7) | 33 (1.9) |
| Leukemia | 1 (5.0) | 1 (4.0) | 3 (1.3) | 2 (2.5) | 4 (3.2) | 2 (2.0) | 8 (2.8) | 5 (2.6) |
| Lymphoma | 5 (2.8) | 5 (1.6) | 4 (4.8) | 3 (2.7) | 3 (2.7) | 1 (1.0) | 12 (3.4) | 9 (1.9) |
| AIDS* | 0 | 0 | 1 + 7 (2.0) | 0 + 8 (-) | 0 + 9 (-) | 0 + 9 (-) | 1 + 16 (2.0) | 0 + 17 (-) |
| Alcohol Consumption | 3 (1.3) | 1 (1.0) | 5 (2.4) | 1 (2.0) | 6 (3.3) | 1 (1.0) | 14 (2.6) | 3 (1.3) |
| Tobacco consumption | 12 (1.7) | 6 (1.5) | 9 (1.8) | 8 (1.1) | 14 (1.9) | 4 (1.0) | 35 (1.8) | 18 (1.2) |
| Total | 48 (11.1) | 40 (5.2) | 42 (7.7) | 37 (2.8) | 44 (11.4) | 35 (2.9) | 134 (10.1) | 112 (3.7) |

\* For those conditions, the number of upsampled notes is also shown *in italics*

Table 2 shows stay-level performances of he **NLP-ML-CLINICAL** pipeline. Outpatient and inpatient cohorts were merged here to yield a higher support for each condition, however split performances are available on Supplementary Table S7. Similarly, per-study performances are available on Supplementary Table S8. Overall, the pipeline reached a macro average F1-score, PPV, sensitivity, specificity of 95.6, 95.3, 95.9 and 99.2, respectively. Additionally, entity-level metrics for this pipeline are available on Supplementary Table S5. Comparing stay-level metrics with entity-level metrics shows the importance of the aggregation algorithm, since it improved macro average F1-score, PPV and sensitivity by 4.7, 2.4 and 6.7 and points respectively. The performance of the isolated qualification algorithm was assessed in Supplementary Table S6, also showing the superiority of the ML approach.

**Table 2:**
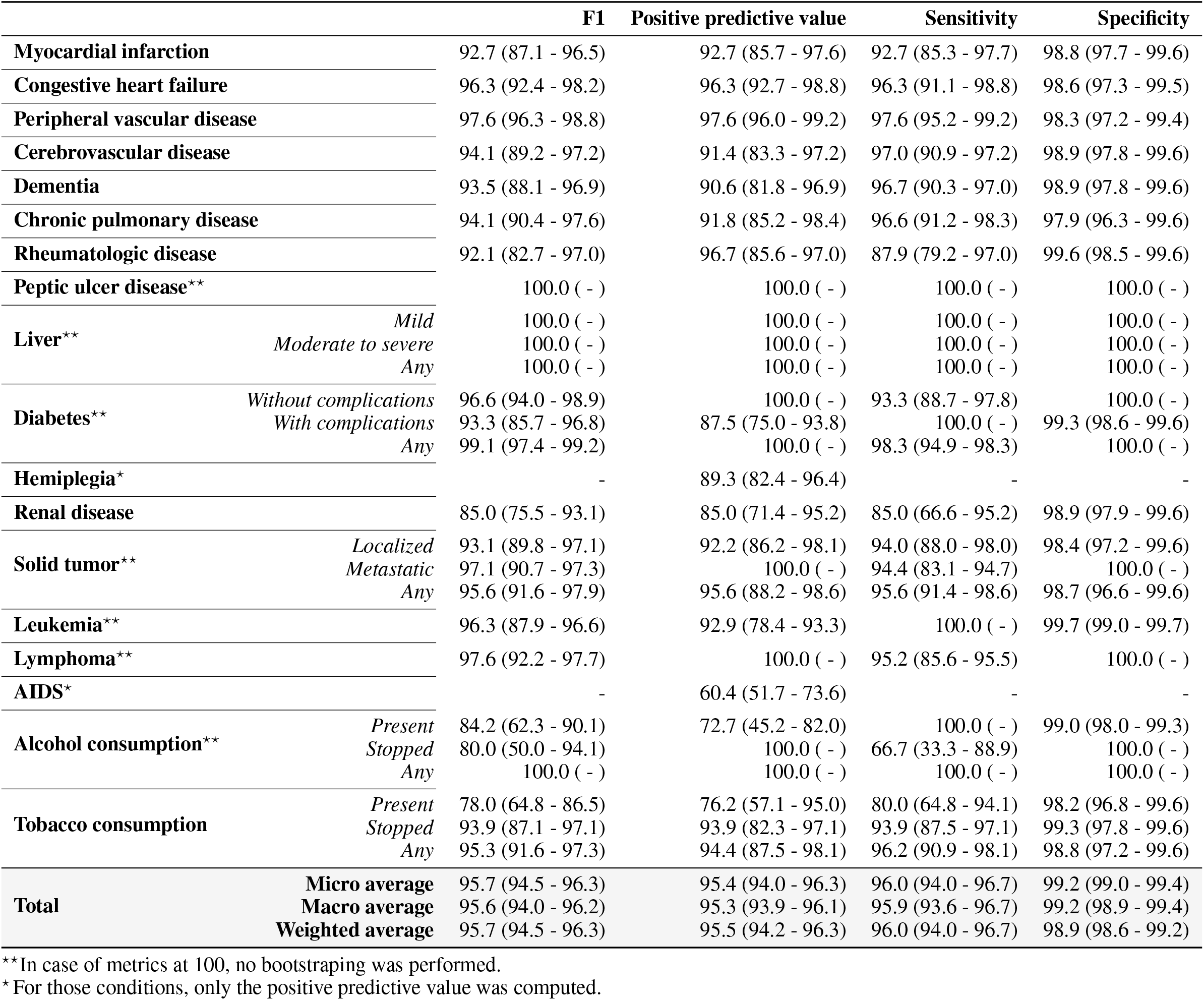
Stay-level performances (F1-score, positive predictive value, sensitivity and specificity) of the main **NLP-ML-CLINICAL** pipeline used to predict 18 conditions from clinical notes considering inpatient and outpatient stays of the three disease-specific cohorts. 95% confidence intervals were estimated by bootstrapping.

|  |  | **F1** | **Positive predictive value** | **Sensitivity** | **Specificity** |
| --- | --- | --- | --- | --- | --- |
| **Myocardial infarction** |  | 92.7 (87.1 - 96.5) | 92.7 (85.7 - 97.6) | 92.7 (85.3 - 97.7) | 98.8 (97.7 - 99.6) |
| **Congestive heart failure** |  | 96.3 (92.4 - 98.2) | 96.3 (92.7 - 98.8) | 96.3 (91.1 - 98.8) | 98.6 (97.3 - 99.5) |
| **Peripheral vascular disease** |  | 97.6 (96.3 - 98.8) | 97.6 (96.0 - 99.2) | 97.6 (95.2 - 99.2) | 98.3 (97.2 - 99.4) |
| **Cerebrovascular disease** |  | 94.1 (89.2 - 97.2) | 91.4 (83.3 - 97.2) | 97.0 (90.9 - 97.2) | 98.9 (97.8 - 99.6) |
| **Dementia** |  | 93.5 (88.1 - 96.9) | 90.6 (81.8 - 96.9) | 96.7 (90.3 - 97.0) | 98.9 (97.8 - 99.6) |
| **Chronic pulmonary disease** |  | 94.1 (90.4 - 97.6) | 91.8 (85.2 - 98.4) | 96.6 (91.2 - 98.3) | 97.9 (96.3 - 99.6) |
| **Rheumatologic disease** |  | 92.1 (82.7 - 97.0) | 96.7 (85.6 - 97.0) | 87.9 (79.2 - 97.0) | 99.6 (98.5 - 99.6) |
| **Peptic ulcer disease\*\*** |  | 100.0 ( - ) | 100.0 ( - ) | 100.0 ( - ) | 100.0 ( - ) |
| **Liver\*\*** | *Mild* | 100.0 ( - ) | 100.0 ( - ) | 100.0 ( - ) | 100.0 ( - ) |
|  | *Moderate to severe* | 100.0 ( - ) | 100.0 ( - ) | 100.0 ( - ) | 100.0 ( - ) |
|  | *Any* | 100.0 ( - ) | 100.0 ( - ) | 100.0 ( - ) | 100.0 ( - ) |
| **Diabetes\*\*** | *Without complications* | 96.6 (94.0 - 98.9) | 100.0 ( - ) | 93.3 (88.7 - 97.8) | 100.0 ( - ) |
|  | *With complications* | 93.3 (85.7 - 96.8) | 87.5 (75.0 - 93.8) | 100.0 ( - ) | 99.3 (98.6 - 99.6) |
|  | *Any* | 99.1 (97.4 - 99.2) | 100.0 ( - ) | 98.3 (94.9 - 98.3) | 100.0 ( - ) |
| **Hemiplegia\*** |  | - | 89.3 (82.4 - 96.4) | - | - |
| **Renal disease** |  | 85.0 (75.5 - 93.1) | 85.0 (71.4 - 95.2) | 85.0 (66.6 - 95.2) | 98.9 (97.9 - 99.6) |
| **Solid tumor\*\*** | *Localized* | 93.1 (89.8 - 97.1) | 92.2 (86.2 - 98.1) | 94.0 (88.0 - 98.0) | 98.4 (97.2 - 99.6) |
|  | *Metastatic* | 97.1 (90.7 - 97.3) | 100.0 ( - ) | 94.4 (83.1 - 94.7) | 100.0 ( - ) |
|  | *Any* | 95.6 (91.6 - 97.9) | 95.6 (88.2 - 98.6) | 95.6 (91.4 - 98.6) | 98.7 (96.6 - 99.6) |
| **Leukemia\*\*** |  | 96.3 (87.9 - 96.6) | 92.9 (78.4 - 93.3) | 100.0 ( - ) | 99.7 (99.0 - 99.7) |
| **Lymphoma\*\*** |  | 97.6 (92.2 - 97.7) | 100.0 ( - ) | 95.2 (85.6 - 95.5) | 100.0 ( - ) |
| **AIDS\*** |  | - | 60.4 (51.7 - 73.6) | - | - |
| **Alcohol consumption\*\*** | *Present* | 84.2 (62.3 - 90.1) | 72.7 (45.2 - 82.0) | 100.0 ( - ) | 99.0 (98.0 - 99.3) |
|  | *Stopped* | 80.0 (50.0 - 94.1) | 100.0 ( - ) | 66.7 (33.3 - 88.9) | 100.0 ( - ) |
|  | *Any* | 100.0 ( - ) | 100.0 ( - ) | 100.0 ( - ) | 100.0 ( - ) |
| **Tobacco consumption** | *Present* | 78.0 (64.8 - 86.5) | 76.2 (57.1 - 95.0) | 80.0 (64.8 - 94.1) | 98.2 (96.8 - 99.6) |
|  | *Stopped* | 93.9 (87.1 - 97.1) | 93.9 (82.3 - 97.1) | 93.9 (87.5 - 97.1) | 99.3 (97.8 - 99.6) |
|  | *Any* | 95.3 (91.6 - 97.3) | 94.4 (87.5 - 98.1) | 96.2 (90.9 - 98.1) | 98.8 (97.2 - 99.6) |
| **Total** | **Micro average** | 95.7 (94.5 - 96.3) | 95.4 (94.0 - 96.3) | 96.0 (94.0 - 96.7) | 99.2 (99.0 - 99.4) |
|  | **Macro average** | 95.6 (94.0 - 96.2) | 95.3 (93.9 - 96.1) | 95.9 (93.6 - 96.7) | 99.2 (98.9 - 99.4) |
|  | **Weighted average** | 95.7 (94.5 - 96.3) | 95.5 (94.2 - 96.3) | 96.0 (94.0 - 96.7) | 98.9 (98.6 - 99.2) |

\*\*In case of metrics at 100, no bootstrapping was performed.
\* For those conditions, only the positive predictive value was computed.

Table 3 compares the stay-level performances of the **NLP-ML-CLINICAL** pipeline with our three alternative pipelines, namely **NLP-ML-GENERIC, NLP-ML-RB** and **CLAIM**, considering only inpatient stays. Overall, the F1-score, PPV and sensitivity were the highest for the **NLP-ML-CLINICAL** pipeline whereas the **CLAIM** pipeline showed the highest specificity.

**Table 3:**
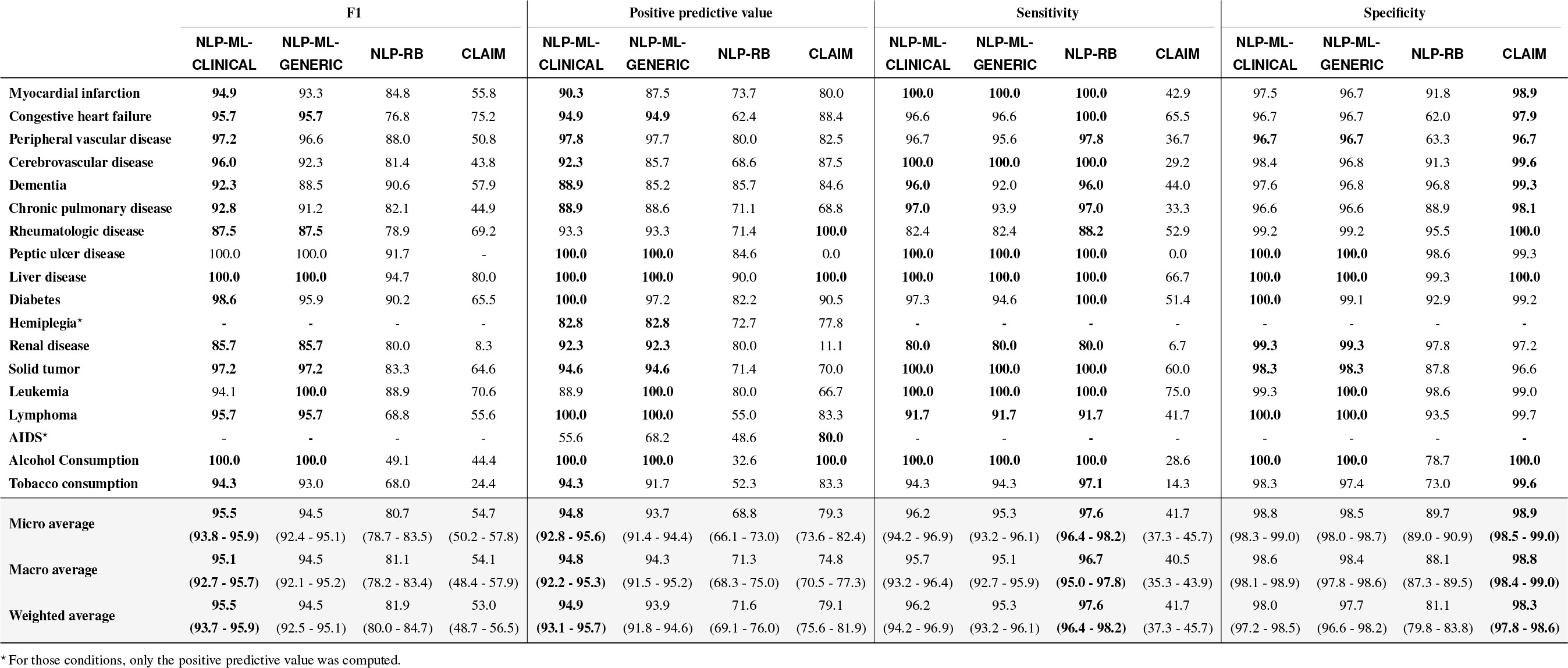
Stay-level performances of the main **NLP-ML-CLINICAL** pipeline compared to three alternative pipelines based on alternative technologies **NLP-ML-GENERIC, NLP-RB** and **CLAIM**. Performances are computed on the inpatient stays of the three cohorts. The 95% confidence intervals of the averaged metrics were computed by bootstrapping.

The prevalence of each condition computed on this same cohort by either the **NLP-ML-CLINICAL** and **CLAIM** pipeline was compared to the chart review prevalence (see Figure 2). Prevalences computed with **NLP-ML-CLINICAL** and **CLAIM** differed from chart review prevalences by 0.82 points and 8.1 points on average, respectively.

**Figure 2:**
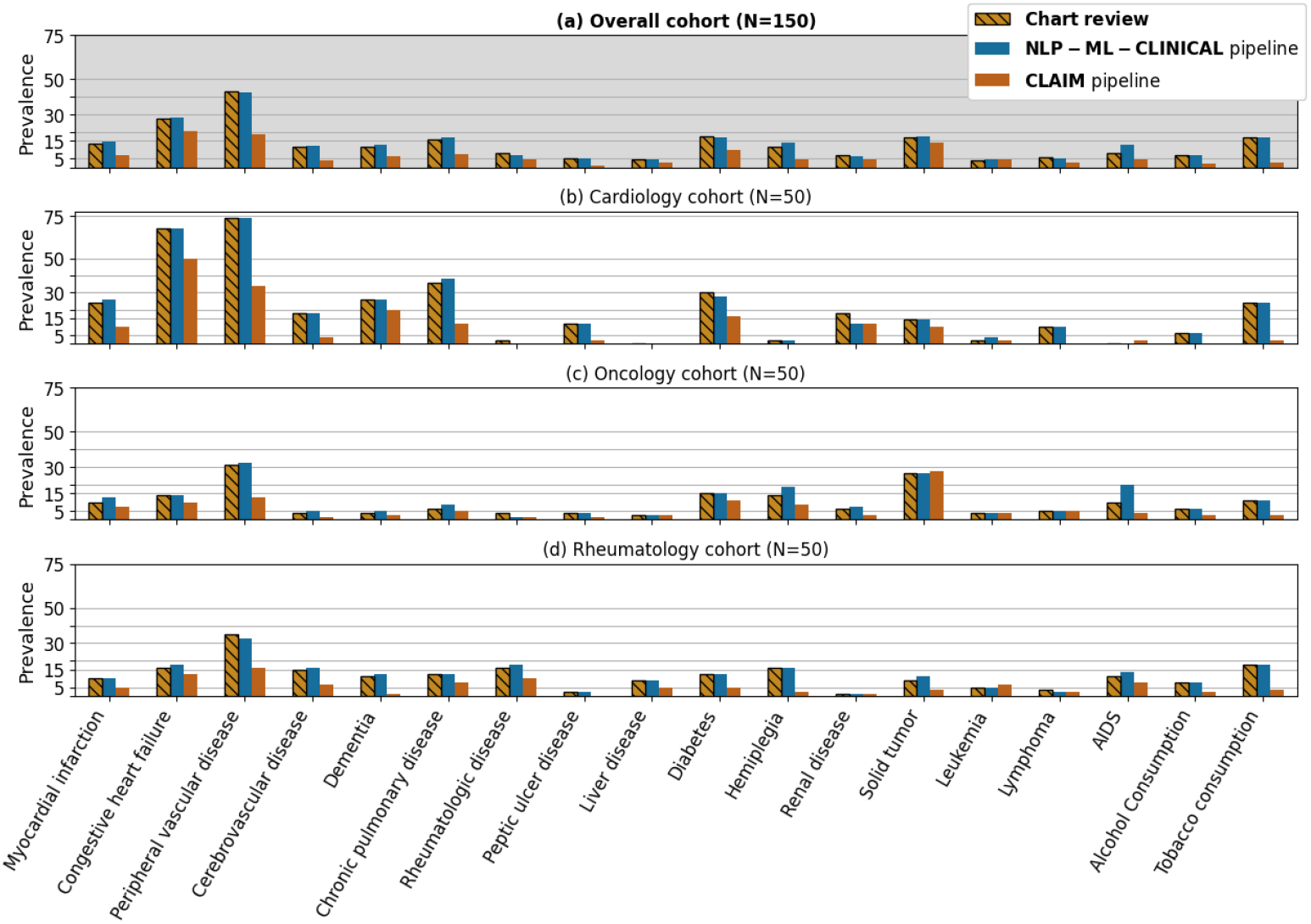
Prevalence of conditions estimated either via the main **NLP-ML-CLINICAL** pipeline (blue), the alternative **CLAIM** pipeline (orange) or a **chart review** of clinical notes (hashed yellow) considering inpatient stays of the cardiology (b), the oncology (c), the rheumatology (d) and the overall (a) cohorts, respectively (number of stays in brackets).

Figure 3 compares different collaborative settings, showing the evolution of the F1-score when training and validating either **NLP-ML-CLINICAL** (left) or **NLP-ML-GENERIC** (right) on each (training cohort, validation cohort) couple. In each of these two cases, the first column shows the F1-score of the pipeline trained on the overall cohort, and the principal diagonal shows the F1-score of the pipeline in a hypothetical less collaborative setting, limiting both the training and validation data to each study’s cohort. In accordance with Table 3, in 75% of all cases **NLP-ML-CLINICAL** was better than **NLP-ML-GENERIC**, thus showing an average improvement related to re-using a clinical language model shared through a secure model hub. Regarding **NLP-ML-CLINICAL**, training on the overall cohort outperformed training on any other study cohort, showing the improvement provided by increasing the collaboration among the three cohort studies to the annotation of a common training dataset. When validating on a specific cohort, results vary and either the pipeline trained on the same cohort as the validation cohort (oncology study) or the pipeline trained on the overall cohort (cardiology and rheumatology cohort) is best performing.

**Figure 3:**
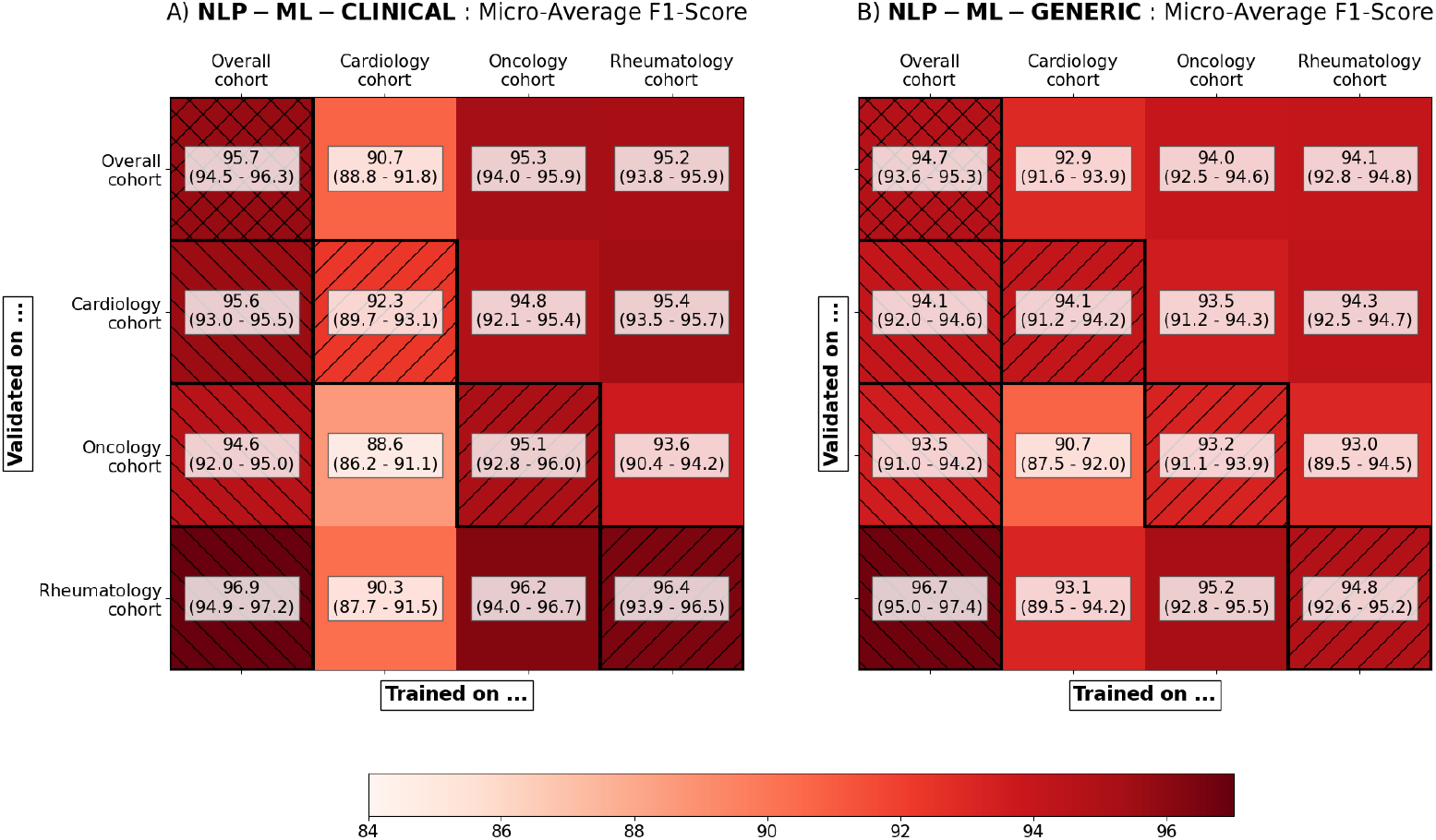
Comparison of the stay-level micro-averaged F1-score of the pipelines and their 95% confidence intervals simulating various levels of collaboration. (A) the main **NLP-ML-CLINICAL** pipeline that reuses the clinical language model of a former project is compared to (B) the **NLP-ML-GENERIC** pipeline that uses only a public language model. In each case, the performances are estimated on every (training study, validation study) couple available. The first column (left hatched) shows performances of the final pipeline trained on the three cohorts together. The main diagonal (right hatched) shows performances of the final pipeline in a hypothetical non-collaborative setting, i.e., trained and validated on the same cohort. 95% confidence intervals were estimated by bootstrapping.

Supplementary Figure S5 compares the CCI computed using **NLP-ML-CLINICAL** with either the **CLAIM** pipeline or CCIs computed at the point of care. Strong discrepancies are observed.

## 4 DISCUSSION

We developed, validated and shared 18 hybrid NLP pipelines to efficiently detect conditions mentioned in clinical notes written in French. We quantified the added value of NLP -compared to claim-based-algorithms, and of advanced-compared to simpler-NLP technologies. Moreover, we measured the performance increase obtained following a collaborative setting that leveraged the hybrid architecture of the pipelines to enable associating various medical experts while complying to data access restrictions enforced on a CDW.

The performances of our main **NLP-ML-CLINICAL** pipeline are comparable to the best published results regarding similar algorithms although slightly lower than those reached by *Singh et al*. ^[13,15,36]^ We emphasize that these studies remain difficult to compare as they focus on different languages (French vs. English) and texts (e.g., *Singh et al*. consider only medical and surgical history sections). Aggregating extractions from the entity level to the stay level significantly improves performances. Notably, sensitivity is greatly improved as the aggregation step allows for missed entities of a specific condition to be compensated by other occurrences of the same condition. Similarly, conditions with two levels of severity benefit from this aggregation since severity would not necessarily be mentioned on each entity. For instance, the diabetes of a patient will be mentioned as complicated, e.g., in the initial section of the clinical notes, and then referred to simply as the patient’s diabetes. Thus, considering entire notes without restrictions to specific sections proved to be beneficial.

Some conditions proved to be more challenging to extract, either because they are mentioned in text with great variety, or because they ask for an additional disambiguation task. We provide a few examples hereby: correctly classifying the current status of alcohol or tobacco consumption -current or stopped-with a rule-based approach was hindered by the highly diverse formulations. The stage of renal failure was often not explicitly mentioned but could be implicit from the context or inferred from other information in the clinical note. AIDS was sometimes mentioned *as is* in the text, but most often it was mentioned as HIV+ along with an opportunistic disease, making it harder to discriminate. Heart failure could often refer to an acute symptom and were difficult to distinguish from a chronic problem.

In accordance with the literature, the superiority of **NLP-ML-CLINICAL** over **NLP-ML-GENERIC** supports the interest of using large language models trained on clinical corpora although their privacy-sensitive nature makes them more difficult to share.^[21,37]^ More generally, it shows the superiority of ML-based pipelines over simpler rule-based and claim-based pipelines, the latter especially struggling with a poor sensitivity. In fact, we showed that hybrid pipelines could both outperform claim-based pipelines when relevant (e.g., on inpatient stays) and replace it when no claim data is available (e.g., on outpatient stays). Claim-based pipeline still outperforms each NLP pipeline regarding specificity: claim data suffers from systemic undercoding, especially with conditions that have little to no impact on the stay’s billing such as alcohol and tobacco consumption, hypertension or asthma. This implies a low sensitivity, but jointly a high specificity.

We demonstrated the relevance of developing common algorithms that jointly support various disease-specific studies conducted on routine data. In fact, both overall performance and generalizability were increased by the collaborative setting: as this setting allows for a larger training set along with more diverse training examples, these gains were expected. Additionally, what couldn’t be measured directly but certainly had a significant impact was the importance of collaboration on the rule-based NER algorithms: concertation meetings showed the complexity of building a robust and clinically relevant terminology, and working with multiple clinicians from various specialties most certainly helped converging toward this goal.

Our work reflects the evolution of privacy protection. Historically, datasets were minimized and de-identified and, once deemed non-sensitive, released to their end users for analysis. Many studies have demonstrated that this solution was no longer applicable in the era of artificial intelligence (AI), as those algorithms often require accessing unminimized and high-dimensional datasets that cannot be anonymized.^[4,20,38–40]^ To address this issue, many call for a switch from an *anonymize-and-release* to a *privacy-through-security* paradigm.^[39,41]^ Secured platforms such as CDWs should host data and models that remain sensitive, while finely controlling accesses and queries. Many different technologies could be employed to this end but, as none is a silver bullet, obtaining privacy-conscientious workflows still requires combining them in architectures adapted to specific usages.^[3]^ We combined access-control, data minimization and model securing by i) leveraging specific opportunities offered by hybrid pipelines and ii) using a secure hub with a dual purpose of facilitating the reuse of pre-trained pipelines and limiting the exposure of privacy-sensitive models.

Our study has limitations. First, inter-annotator agreement could not be measured as each investigator accessed only her disease-specific cohort. Also, using a pre-annotation step before manual annotation might lead to a confirmation bias.^[42]^ Nevertheless, it substantially speeds up the annotation and allows to reach higher volumes of annotated data, which arguably increases the robustness of the process. Second, although different technologies were explored, we focused for this comparison on the qualification algorithm and always used a simple aggregation rule to define variables at the stay level. Implementing more advanced aggregation algorithms may improve the algorithms’ performances.

We also separately analyzed either clinical notes or claim data: jointly considering text and additional variables such as laboratory test results, drugs or claim data may improve the performances.^[7]^ The ML-based qualification algorithm could also be extended to classify, if relevant, the status of the condition, since this task can be complex and highly dependent on context. Third, the privacy-preserving workflow we propose is a single step in the direction of efficient and privacy-conscientious architectures as i) data exposition, although reduced, remains non-negligible, ii) this workflow does not apply to other tasks such as data exploration that is a cornerstone of data analysis, and iii) it relies on the prior integration of data in a common technical and regulatory platform such as a CDW and consequently does not address the central issue of model sharing across organizations.

## 5 CONCLUSION

We followed a new collaborative and privacy preserving workflow to develop and validate 18 hybrid rule-based/ML pipelines dedicated to the automatic detection of common conditions in clinical notes written in French. Thus, we demonstrated that using advanced NLP technologies led to higher performances when compared to classical approaches and that hybrid architectures could be leveraged to associate various experts working on a common CDW. This work is a first step in the direction of better adapting data analysis workflows both to emerging ML technologies and to the regulatory constraints. We hope that this work will pave the way to closer collaborations among a wide community of clinicians, investigators and engineers working on CDWs.

## Supporting information

RECORD Checklist

ICD10 Codes

## Data Availability

Models and pipelines codes are available online at https://github.com/aphp-datascience/study-collaborative-workflow-nlp

## 6 ACKNOWLEDGEMENTS

We thank the clinical data warehouse (Entrepôt de Données de Santé, EDS) of the Greater Paris University Hospitals for its support and the realization of data management and data curation tasks. We thank Cyril Charron and Quentin Delannoy for fruitful discussions.

## 7 CODE AVAILABILITY

The code for the hybrid model and the experiments of this study were made available on a dedicated GitHub repository.^[43]^ As mentionned in the study, the NER algorithms were made available in the 0.8.1 release of the EDSNLP library and the trained qualification algorithm wa made available into APHP’s CDW through a secure registry.^[34]^

## 10 AUTHOR CONTRIBUTIONS STATEMENT

T.P.-J., C.G., E.K., E.B., R.B. had access to the data in the study. They take responsibility for the integrity of the data and the accuracy of the data analysis. All authors interpreted data and made critical intellectual revisions of the manuscript.

Concept and design: T.P.-J., C.G., E.K., E.B., X.T., G.C., M.F., R.B.

Annotation and interpretation of data: C.G., E.K., E.B.

Manuscript drafting: T. P.-J., R.B.

T. P.-J. and R.B. did the literature review.

Algorithm development: T.P.-J.

Supervision: R.B.

## 11 SUPPLEMENTARY MATERIAL

Supplementary material is available online on *MedRvix*.

## REFERENCES

[1] Eric J Topol. High-performance medicine: the convergence of human and artificial intelligence. Nature medicine, 25(1):44–56, 2019.

[2] Michael Moor, Oishi Banerjee, Zahra Shakeri Hossein Abad, et al. Foundation models for generalist medical artificial intelligence. Nature, 616(7956):259–265, 2023.

[3] National Science and Technology Concil. National strategy to advance privacy-preserving data sharing and analytics. https://www.whitehouse.gov/wp-content/uploads/2023/03/National-Strategy-to-Advance-Privacy-Preserving-Data-Sharing-and-Analytics.pdf. Accessed: 20-7-2023.

[4] Eric Lehman, Evan Hernandez, Diwakar Mahajan, et al. Do we still need clinical language models? arXiv preprint arXiv:2302.08091, 2023.

[5] Nicholas Carlini, Florian Tramer, Eric Wallace, et al. Extracting training data from large language models. In 30th USENIX Security Symposium (USENIX Security 21), pages 2633–2650, 2021.

[6] Douglas G Manuel, Laura C Rosella, and Thérése A Stukel. Importance of accurately identifying disease in studies using electronic health records. Bmj, 341, 2010.

[7] Seyedmostafa Sheikhalishahi, Riccardo Miotto, Joel T Dudley, et al. Natural language processing of clinical notes on chronic diseases: systematic review. JMIR medical informatics, 7(2):e12239, 2019.

[8] Alexandre Lampros, Camille Montardi, Louis Journeau, et al. Association des comorbidités psychiatriques avec la durée de séjour des patients en médecine interne d’aval des urgences. La Revue de Médecine Interne, 41(6):360–367, 2020.

[9] Mary E Charlson, Peter Pompei, Kathy L Ales, and C Ronald MacKenzie. A new method of classifying prognostic comorbidity in longitudinal studies: development and validation. Journal of chronic diseases, 40(5):373–383, 1987.

[10] Richard A Deyo, Daniel C Cherkin, and Marcia A Ciol. Adapting a clinical comorbidity index for use with icd-9-cm administrative databases. Journal of clinical epidemiology, 45(6):613–619, 1992.

[11] Vijaya Sundararajan, Toni Henderson, Catherine Perry, et al. New icd-10 version of the charlson comorbidity index predicted in-hospital mortality. Journal of clinical epidemiology, 57(12):1288–1294, 2004.

[12] Jen-Hsiang Chuang, Carol Friedman, and George Hripcsak. A comparison of the charlson comorbidities derived from medical language processing and administrative data. In Proceedings of the AMIA Symposium, page 160. American Medical Informatics Association, 2002.

[13] Balwinder Singh, Amandeep Singh, Adil Ahmed, et al. Derivation and validation of automated electronic search strategies to extract charlson comorbidities from electronic medical records. Mayo Clinic Proceedings, 87(9):817–824, September 2012.

[14] Hojjat Salmasian, Daniel E Freedberg, and Carol Friedman. Deriving comorbidities from medical records using natural language processing. Journal of the American Medical Informatics Association, 20(e2):e239–e242, 2013.

[15] Adam N Berman, David W Biery, Curtis Ginder, et al. Natural language processing for the assessment of cardiovascular disease comorbidities: The cardio-canary comorbidity project. Clinical Cardiology, 44(9):1296–1304, 2021.

[16] Seungwon Lee, Chelsea Doktorchik, Elliot Asher Martin, et al. Electronic medical record–based case phenotyping for the charlson conditions: Scoping review. JMIR medical informatics, 9(2):e23934, 2021.

[17] Alexander Turchin and Luisa F Florez Builes. Using natural language processing to measure and improve quality of diabetes care: a systematic review. Journal of Diabetes Science and Technology, 15(3):553–560, 2021.

[18] Henrique Dias Pereira dos Santos, Ana Helena DPS Ulbrich, Vinicius Woloszyn, and Renata Vieira. An initial investigation of the charlson comorbidity index regression based on clinical notes. In 2018 IEEE 31st International Symposium on Computer-Based Medical Systems (CBMS), pages 6–11. IEEE, 2018.

[19] Le Zheng, Yue Wang, Shiying Hao, et al. Web-based real-time case finding for the population health management of patients with diabetes mellitus: A prospective validation of the natural language processing–based algorithm with statewide electronic medical records. JMIR medical informatics, 4(4):e6328, 2016.

[20] Basile Dura, Charline Jean, Xavier Tannier, et al. Learning structures of the french clinical language: development and validation of word embedding models using 21 million clinical reports from electronic health records. arXiv preprint arXiv:2207.12940, 2022.

[21] Jinhyuk Lee, Wonjin Yoon, Sungdong Kim, et al. Biobert: a pre-trained biomedical language representation model for biomedical text mining. Bioinformatics, 36(4):1234–1240, 2020.

[22] Elizabeth Ford, John A Carroll, Helen E Smith, et al. Extracting information from the text of electronic medical records to improve case detection: a systematic review. Journal of the American Medical Informatics Association, 23(5):1007–1015, 2016.

[23] Yanshan Wang, Liwei Wang, Majid Rastegar-Mojarad, et al. Clinical information extraction applications: a literature review. Journal of biomedical informatics, 77:34–49, 2018.

[24] Zeljko Kraljevic, Thomas Searle, Anthony Shek, et al. Multi-domain clinical natural language processing with medcat: the medical concept annotation toolkit. Artificial intelligence in medicine, 117:102083, 2021.

[25] Philip John Gorinski, Honghan Wu, Claire Grover, et al. Named entity recognition for electronic health records: a comparison of rule-based and machine learning approaches. arXiv preprint arXiv:1903.03985, 2019.

[26] Jordan Jouffroy, Sarah F Feldman, Ivan Lerner, et al. Hybrid deep learning for medication-related information extraction from clinical texts in french: Medext algorithm development study. JMIR medical informatics, 9(3):e17934, 2021.

[27] Aurélie Névéol, Hercules Dalianis, Sumithra Velupillai, et al. Clinical natural language processing in languages other than english: opportunities and challenges. Journal of biomedical semantics, 9(1):1–13, 2018.

[28] Rebecca Knowles, Bilal A Mateen, and Yo Yehudi. We need to talk about the lack of investment in digital research infrastructure. Nature Computational Science, 1(3):169–171, 2021.

[29] Nicholas Carlini, Chang Liu, U. lfar Erlingsson, et al. The secret sharer: Evaluating and testing unintended memorization in neural networks. In 28th USENIX Security Symposium (USENIX Security 19), pages 267–284, 2019.

[30] The European Parliament and the Concil of the European Union. Regulation (eu) 2016/679. https://eur-lex.europa.eu/eli/reg/2016/679/oj. Accessed: 20-7-2023.

[31] Eric I Benchimol, Liam Smeeth, Astrid Guttmann, et al. The reporting of studies conducted using observational routinely-collected health data (record) statement. PLoS medicine, 12(10):e1001885, 2015.

[32] Romain Bey, Romain Goussault, François Grolleau, et al. Fold-stratified cross-validation for unbiased and privacy-preserving federated learning. Journal of the American Medical Informatics Association, 27(8):1244–1251, 2020.

[33] Shawn N Murphy, Griffin Weber, Michael Mendis, et al. Serving the enterprise and beyond with informatics for integrating biology and the bedside (i2b2). Journal of the American Medical Informatics Association, 17(2):124–130, 2010.

[34] Basile Dura, Perceval Wajsburt, Thomas Petit-Jean, et al. EDS-NLP: efficient information extraction from French clinical notes, July 2023.

[35] Louis Martin, Benjamin Muller, Pedro Javier Ortiz Suárez, et al. Camembert: a tasty french language model. arXiv preprint arXiv:1911.03894, 2019.

[36] Anthony Shek, Zhilin Jiang, James Teo, et al. Machine learning-enabled multitrust audit of stroke comorbidities using natural language processing. European Journal of Neurology, 28(12):4090–4097, 2021.

[37] Yanis Labrak, Adrien Bazoge, Richard Dufour, et al. Drbert: A robust pre-trained model in french for biomedical and clinical domains. bioRxiv, pages 2023–04, 2023.

[38] Charu C Aggarwal. On k-anonymity and the curse of dimensionality. In VLDB, volume 5, pages 901–909, 2005.

[39] Yves-Alexandre de Montjoye, Ali Farzanehfar, Julien Hendrickx, and Luc Rocher. Solving artificial intelligence’s privacy problem. Field Actions Science Reports. The journal of field actions, Special Issue 17:80–83, 2017.

[40] Luc Rocher, Julien M Hendrickx, and Yves-Alexandre De Montjoye. Estimating the success of re-identifications in incomplete datasets using generative models. Nature communications, 10(1):1–9, 2019.

[41] Yves-Alexandre De Montjoye, Sébastien Gambs, Vincent Blondel, et al. On the privacy-conscientious use of mobile phone data. Scientific data, 5(1):1–6, 2018.

[42] Karën Fort and Benôit Sagot. Influence of pre-annotation on pos-tagged corpus development. In The fourth ACL linguistic annotation workshop, pages 56–63, 2010.

[43] Thomas Petit-Jean and Romain Bey. aphp-datascience/study-collaborative-workflow-nlp: v1.0.0, doi: 10.5281/zenodo.8328741, url: 10.5281/zenodo.8328741, August 2023.

