## Supplementary Material for "Collaborative and privacy-preserving workflows on a clinical data warehouse: an example developing natural language processing pipelines to detect medical conditions"

### 1 Appendix A: Description of the cardiology, rheumatology and oncology cohorts

#### 1.1 Per-cohort inclusion criteria

##### 1.1.1 Oncology cohort

The cohort comprises patients with a diagnosis of cancer. Characterization is done via diagnosis claim codes (*International Classification of Diseases, 10^th^ revision* -ICD-10-), clinical procedure codes (French *Classification Commune des Actes Médicaux* -CCAM-) or diagnosis related group codes (French *Groupement Homogène de Malades* -GHM-) attached to a hospital stay occurring between 2006 and 2020 (Supplementary Table S1 and [1]).

| ICD-10 Codes | CCAM Codes | GHM Codes |
|---|---|---|
| C00-C97, D00-D09, D37-D48, B21, D630, E883, G533, G550, G631, G732, G941, J700, J701, K520, K627, L412, L580, L581, L598, L599, M360, M361, M906, M907, M962, M965, N304, O356, T451, T66, T860, Z08, Z510, Z511, Z9480, Y431, Y432, Y433, Y632, Z85, Z923, Z926 | YYYY042, ZZNL065, ZZML001, ZZNL063, FEFF002, ZZML003, ZZNL054, ZZMK018, ZZMP017, ZZNL051, HLNM001, BGLA002, HLNK001, ZZNL050, EDLF017, ZZML002, HKFA007, ZZNL048, AFLB003, ZZNL062, ZZMP015, ZZLF900, EDLF014, EDLF016, ZZMK020, ZZNL053, FDFB001, FELF009, FELF010, GGLB001, ZZNL061, GENE003, HHGE010, KCNL004, ZZMK011, ZZNL058, HELE001, EDLF015, ZZLF004, ZZMP012, ZANL001, ZZNL059, ZZNL064, AGMP001, EDLL002, FEFF001, GGLB008, HLNN900, HPLA002, JDLA001, JDLA001, JLNL005, ZZMK002, ZZMK013, ZZMK024, ZZMP016, ZZNL003, ZZNL005, ZZNL016, ZZNL016, ZZNL018, ZZNL019, ZZNL047, ZZNL060 | 01M261, 01M262, 01M263, 01M264, 01M26T, 03M071, 03M072, 03M073, 03M074, 03M07T, 06C161, 06C162, 06C163, 06C164, 06M051, 06M052, 06M053, 06M054, 06M05T, 06M131, 06M132, 06M133, 06M134, 06M13T, 07C061, 07C062, 07C063, 07C064, 07C091, 07C092, 07C093, 07C094, 07M061, 07M062, 07M063, 07M064, 07M06T, 08C291, 08C292, 08C293, 08C294, 08C29J, 08M241, 08M242, 08M243, 08M244, 08M24T, 09C041, 09C042, 09C043, 09C044, 09C051, 09C052, 09C053, 09C054, 09C05J, 09M101, 09M102, 09M103, 09M104, 09M10T, 10C111, 10C112, 10C113, 10C114, 12C051, 12C052, 12C053, 12C054, 12C091, 12C092, 12C093, 12C094, 12C111, 12C112, 12C113, 12C114, 12M031, 12M032, 12M033, 12M034, 12M03T, 13C051, 13C052, 13C053, 13C054, 13C111, 13C112, 13C113, 13C114, 13C11J, 13C141, 13C142, 13C143, 13C144, 13M031, 13M032, 13M033, 13M034, 13M03T, 17C021, 17C022, 17C023, 17C024, 17C031, 17C032, 17C033, 17C034, 17C03J, 17M051, 17M052, 17M053, 17M054, 17M061, 17M062, 17M063, 17M064, 17M06T, 17M081, 17M082, 17M083, 17M084, 17M08T, 17M091, 17M092, 17M093, 17M094, 17M09T, 17M111, 17M112, 17M113, 17M114, 17M11T, 17M121, 17M122, 17M123, 17M124, 17M12T, 28Z07Z, 28Z10Z, 28Z11Z, 28Z18Z, 28Z19Z, 28Z20Z, 28Z21Z, 28Z22Z, 28Z23Z, 28Z24Z, 28Z25Z, 27Z021, 27Z022, 27Z023, 27Z024, 27Z03Z, 27Z04J |

Table S1: Codes used to select the oncology cohort

##### 1.1.2 Cardiology cohort

The cohort comprises patients aged 75 or more, hospitalized after August 2017 in a cardiology, pneumology or geriatry department and presenting either a diagnosis of acute cardiac failure, pulmonary embolism, pulmonary infection or chronic pulmonary obstructive disease (extraction via diagnosis claim codes), and a dosage of natriuretic peptides. Biological Characterization is done via diagnosis claim codes and laboratory test results (*Logical Observation Identifiers Names and Codes -LOINC-*). See Supplementary Table S2

##### 1.1.3 Rheumatology cohort

The cohort comprises patients aged 15 or older with at least one hospital stay between January 7th, 2017 and December 31st, 2020 with a medical history of either systemic lupus erythematosus, Takayasu disease, scleroderma or antiphos-

| ICD-10 Codes | LOINC Codes |
|---|---|
| I50, I110, I26, J441, J209, J46, J12, J13, J14, J15, J16, J17, J18, J40, J41, J42, J43, J44, J45, J47, J20 | 33762-6, 30934-4 |

Table S2: ICD-10 diagnostic codes and LOINC laboratory codes used to select the cardiology cohort

pholipid syndrome. Characterization is done via diagnosis claim codes and verbatim mentions of these diseases in clinical reports. See Supplementary Table S3

| Disease | ICD-10 Codes | Keyword(s) (*French*) |
|---|---|---|
| Lupus | M310, M321, M328, M329, L930, L931 | ”lupus” |
| Takayasu disease | M314 | ”Takayasu” |
| Sclerodermia | M340, M341, M348, M349 | ”sclerodermie”, ”CREST” |
| Antiphospholipid antibody syndrome | D686 | ”SAPL”, ”syndrome des antiphospholipides”, ”CAPS” |

Table S3: ICD-10 diagnostic codes and keywords used to select the rheumatology cohort.

##### 1.2 Training and validation datasets

To split the datasets of the different projects in three development datasets and three validation datasets while avoiding data leakage between these sub-cohorts, we would ideally need to share patient identities. Yet, identity linkage between the different isolated environments is impossible due to privacy issues. We therefore use the recently published technique of fold-stratified (cross)-validation, a privacy-preserving technique that avoids patient-level linkage while enabling splitting the datasets in development datasets and validation datasets.[2] In order to apply this technique, we stratify the cohorts using a covariate that is shared among pseudonymized records, namely the birth date.

Each cohort was split into three sub-cohorts. As shown on the flowchart (Supplementary Figure S1), we used this number along with a hash function to

1. Stratify patients and avoid multiple annotations of the same visits: to do so, project n°i worked with patients which birthday $BD$ verifies $Hash(BD) \equiv i \pmod 3$ with $Hash$ being the MurmurHash3 function.
2. Further split patients into training sets and a validation set in each project: if $Hash(Hash(BD)) \equiv j \pmod 2$, the corresponding patient would be put into the dataset for the $j$-th iteration. Patients with $j = 0$ were included in the validation dataset.

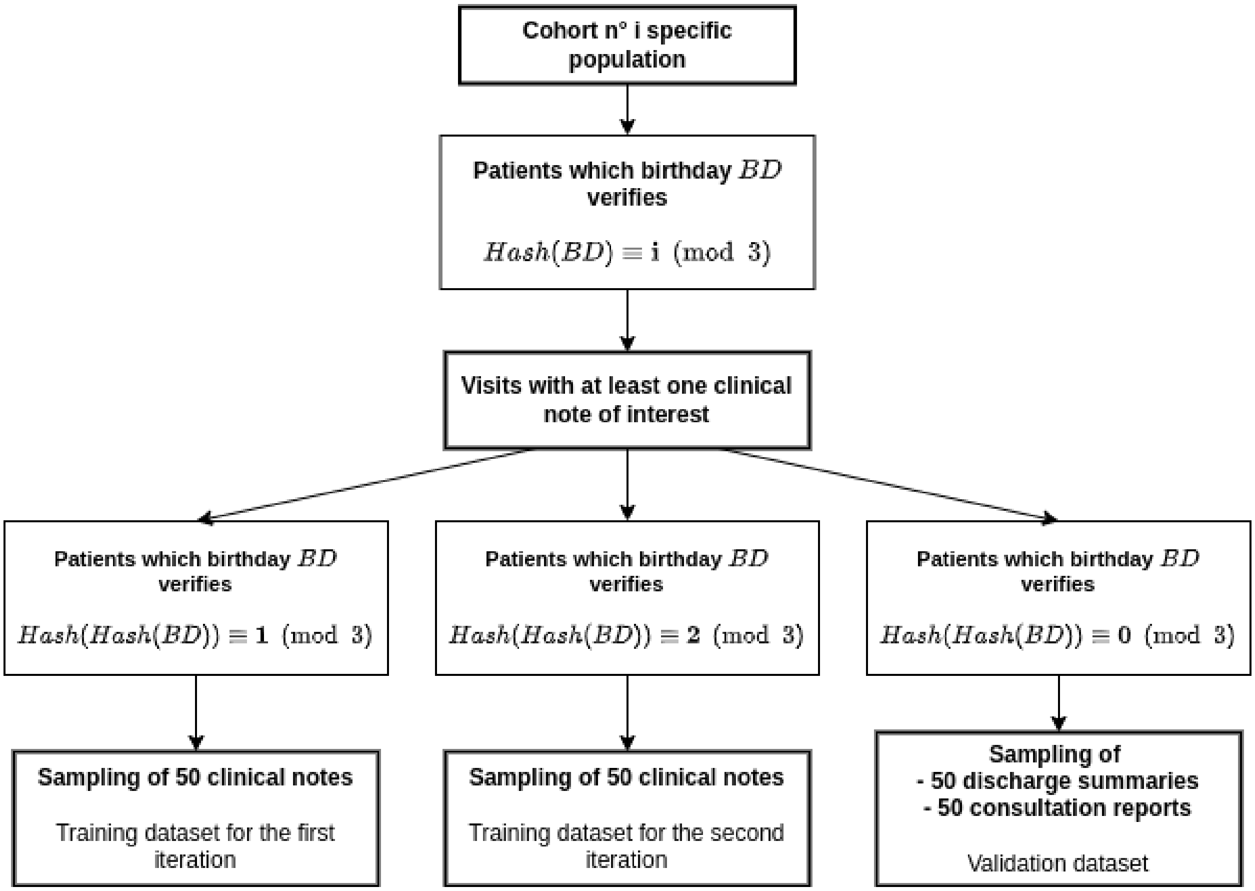


Figure S1: Inclusion flowchart for one study guaranteeing the absence of data leakage among studies

#### 2 Appendix B: Details on the algorithms

##### 2.1 Natural language processing pipeline

The input of the natural language processing (NLP) pipeline consists of a single clinical note. Its structure is schematically shown on Supplementary Figure S2.

###### 2.1.1 Preprocessing

The **preprocessing** module splits the raw text into tokens (individual words) and sentences. It then normalizes the text by

- switching case to lowercase (*Diabète* et *'Behçet'* $\rightarrow$ *diabète* et *'behçet'*)
- removing diacritics (*diabète* et *'behçet'* $\rightarrow$ *diabete* et *'behcet'*)
- uniforming quotes and apostrophes (*diabete* et *'behcet'* $\rightarrow$ *diabete* et *"behcet"*)

Finally, it detects frequent pollution patterns in texts which were identified as potential sources of false positives for the named entity recognition (NER) algorithms. For instance, it will tag e-mail address and websites URLs (e.g., to avoid matching patterns such as `www.diabetes.fr` or ``) or parsed laboratory results, which often contain disease names (moreover, the exploitation of such biology results is outside of this article's scope).

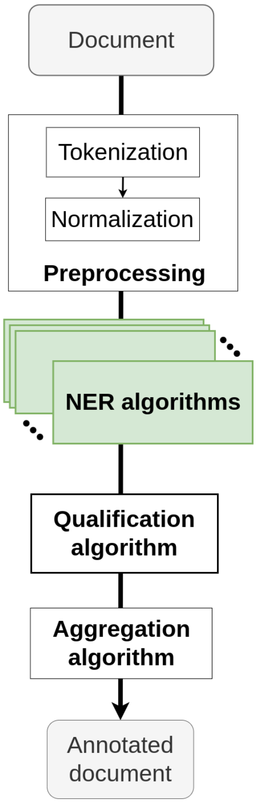


Figure S2: Structure of the natural language processing pipeline. In green are the rule-based, iteratively-improved components (i.e., the Named Entity Recognition algorithms).

##### 2.1.2 NER algorithms

For each comorbidity, the NER algorithm consists of a set of *matching dictionaries*. A *matching dictionary* has three main components which are called sequentially in the algorithm (see Supplementary Figure S3 for an example of NER algorithm structure).

The first component is the *matching component*. Using a set of regular expressions, it will find *anchors* (i.e., raw mentions of a specific disease or condition) in text. The second component is the *exclusion component*. For each *anchor*, it applies exclusion rules on its surrounding window to remove incorrect or undesirable matches. The third component is the *refining component*. Like the *exclusion component*, it will search in each *anchor*'s surrounding window to extract additional information that will help further discriminate the match (for instance to set, if applicable, the status or severity level).

##### 2.1.3 Qualification algorithm

The **qualification algorithm** characterizes the context of each occurrence. The simple detection of an occurrence of a concept in clinical notes is often misleading, as an occurrence of a term may for instance be negated, formulated as a hypothesis or related to a member of the family. Although the paper's main pipeline uses a qualifier based on Machine Learning (ML), both a **rule-based approach** and an **ML-based approach** were carried out.

The rule-based algorithm (Supplementary Figure S4, left) uses a list of regular expression names *cues* to detect marks of negation, hypothesis or familial context. By linking those *cues* to extracted conditions, we flagged each condition as being either

- negated: Is the detected condition negated in the text ?
- family-related: Is the detected condition referring to a member of the patient instead of the patient himself ?
- hypothetical: Is there a hypothesis marker ?

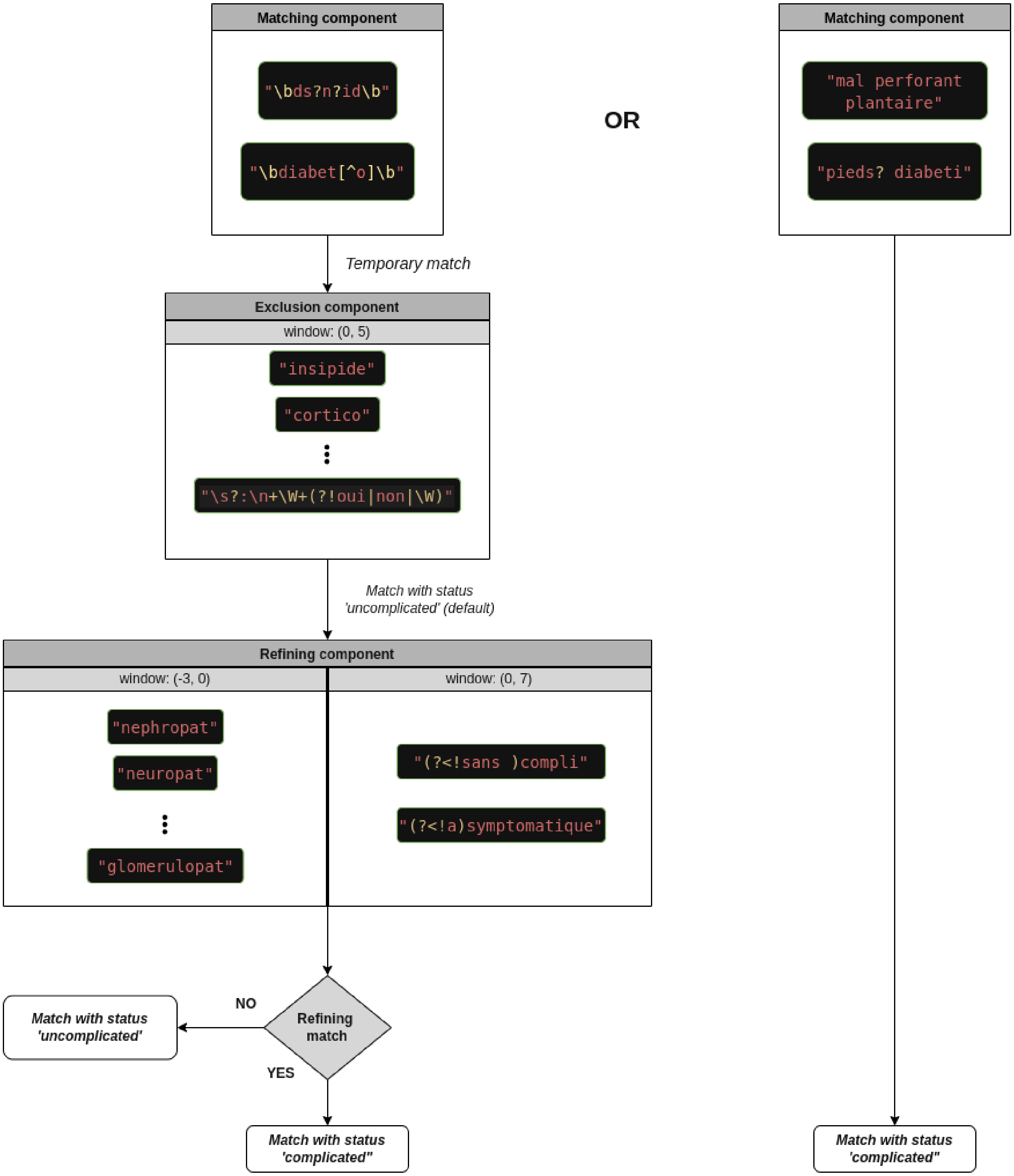


Figure S3: Illustration of a named entity recognition algorithm considering the example of *Diabetes*. Only subsets of patterns are shown in each component.

This algorithm was not developed as part of this study. It was implemented beforehand in the EDS-NLP libray and adapts the NegEx algorithm to the french language.[3,4].

Finally, a unique boolean flag is constructed by taking the boolean OR of those three flags.

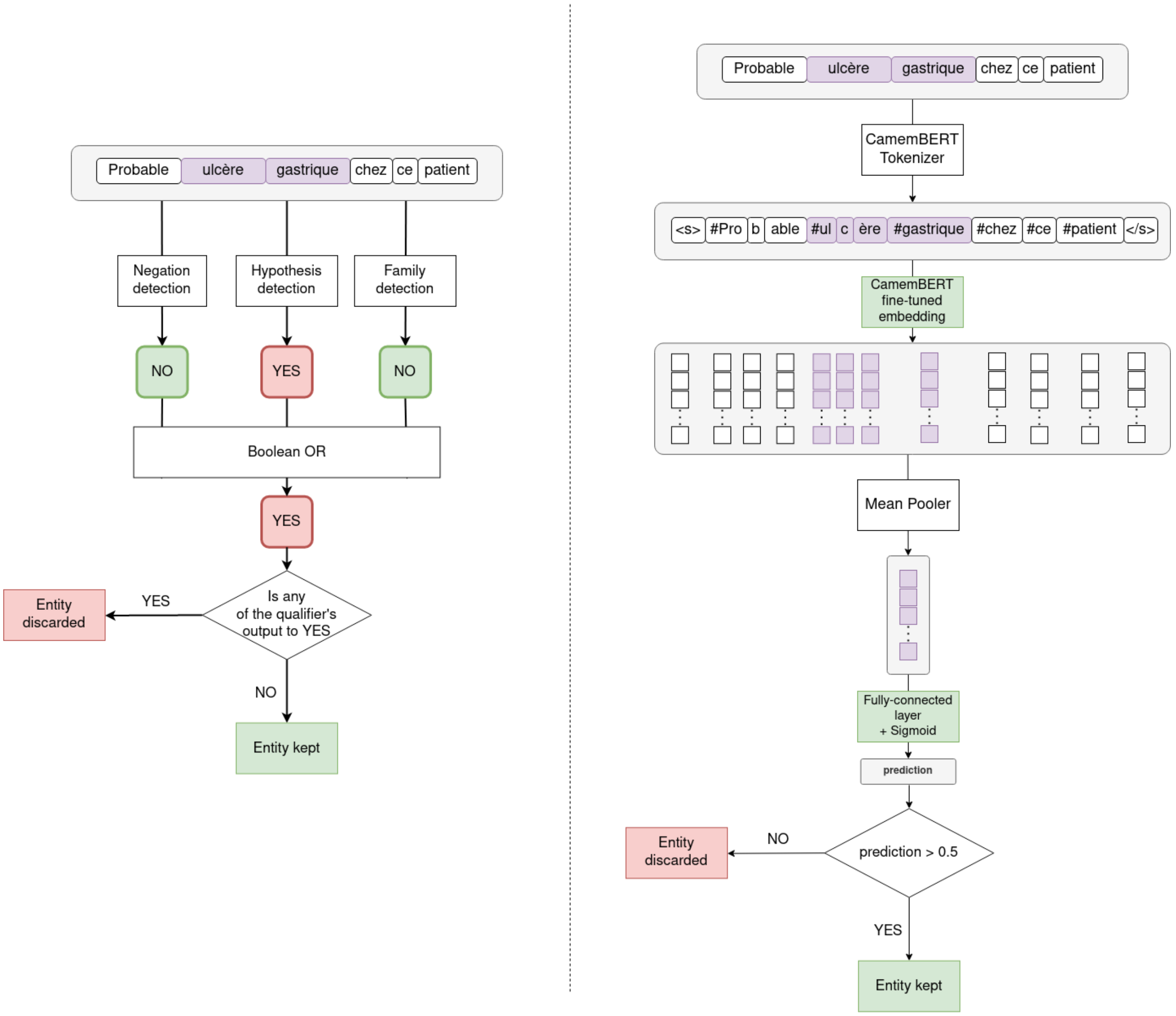


Figure S4: Structure of the rule-based (left) and machine learning (ML)-based (right) qualification algorithms. Regarding the ML-based algorithm, modules whose parameters were tuned during the training procedure are shown in green.

Instead of tagging multiple modalities like the rule-based qualification algorithm, for this ML approach we trained a neural network to classify an extraction as valid or not, encapsulating the qualification in a single boolean variable (Supplementary Figure S4, right). For the classification head, a mean pooler layer was used to average all embedding vectors from an entity into a single vector. Following was a fully connected layer. This architecture allowed us to handle the qualification task as a sentence classification task, assigning one boolean label to each snippet.

##### 2.1.4 Document-level classification

The **document-level classification module** aims at reconciling, for each concept, extractions from a single document by doing a rule-based aggregation. The goal is then to end up with document-level labels. We used a simple but efficient rule that considered, for each condition, the set of all related entities which were not discarded by the qualification module. If this set was not empty, we considered the condition as present. Moreover, in the case of conditions with multiple statuses, we kept the status of the most severe entity from the set.

### 3 Appendix C: Alorithms development

#### 3.1 Documents sampling

We upsampled the documents used to train the ML algorithm in order i) to optimize the annotated dataset by selecting in priority documents with many mentions of the targeted conditions and ii) to select documents that contain the most interesting patterns to learn, for instance those that were not detected by the simple rule-based algorithm. Therefore, prior to any annotation campaign we computed two variables for each document of the training dataset:

- $E$ the number of entities detected by the NER and Qualification algorithms of the simple **NLP-RB** pipeline in the case of discharge summaries and consultation reports.
- $FN$ the number of *probable false negatives* of the **NLP-RB** pipeline, which we computed for inpatient stays only by first applying the **CLAIM** pipeline and then counting the number of conditions detected by the later but not by the **NLP-RB** pipeline. Indeed, as the **CLAIM** pipeline features a low specificity, comparing both leads to a lower bond of the number of false negatives.

In the case of outpatient stays, we used a power distribution ($p(x) = 5x^4, x \in [0, 1]$), a group was selected following the probability law:

$$\mathbb{P}(E) = \left(\frac{E+1}{E_{max}}\right)^5 - \left(\frac{E}{E_{max}}\right)^5$$

with $E_{max}$ the maximal number of entities detected in a document of the dataset. Once the group was selected, a document was chosen among this group using a uniform sampling. This process was repeated until the number of required documents were selected.

In the case of inpatient stays, a first half of the documents were selected following the same method and a second half of the documents was selected to maximize the number of *probable false negatives*. For the selection of the latter, eligible documents were grouped by $FN$ and a group was similarly selected following the probability law:

$$\mathbb{P}(FN) = \left(\frac{FN+1}{FN_{max}}\right)^5 - \left(\frac{FN}{FN_{max}}\right)^5$$

with $FN_{max}$ the maximal number of *probable false negatives* detected in a document of the dataset. Adopting the same method as before, one document was then selected uniformly in the group. To consider equally both aspects we alternatively selected a single document following each method until the required amount of documents was reached.

The exponent four was chosen as it looked relevant after a visual inspection of the selected datasets. We emphasize that this obviously biased upsampling does not affect the reliability of the final performances that were computed using a uniformly sampled dataset.

#### 3.2 De-identification and consolidation of the training dataset

During the annotation process, along with each extraction, a snippet of text is shown to the annotator. This snippet is centered around the extraction (a sentence before and a sentence after the entity). To prepare concertation meetings, which were centered around analysing and discussing incorrect extractions, the annotators were also asked to check this snippet on incorrect extractions to remove any identifying data (e.g., names, places of residence, dates of birth, etc.) and, if applicable, replacing it with a placeholder. For extra caution, if any doubt is emitted by an annotator on the potential re-identification of a patient using the remaining information contained in a snippet, the snippet was discarded.

We advocate for the capacity to centralize the annotated snippets in a development environment accessed only by data scientists/engineers in charge of the model training. Yet, this workflow is not yet validated and we can consequently exchange among environments only highly de-identified data. To be able to emulate the workflow shown in Figure 1, we therefore conducted an a-posteriori time-consuming de-identification of all snippets that relied on two stages. First, entity types listed in Table S10 were manually annotated and automatically replaced in the snippets by a plausible placeholder. Second, when the snippet could lead to indirect identification (e.g., listing exhaustively the patient's family history), it was excluded from the dataset, meaning they could not be used to train the ML-based qualification algorithm.

Due to the iterative structure of the annotation process, some annotations carried out during the first session may not be relevant anymore when reaching the end of the process. For instance, it was decided during the process to exclude *claudications* for the *Peripheral vascular disease* item, or to exclude *occasional* drinking from the *alcohol consumption* item. We also corrected those definition drift during the review mentionned above.

##### 3.3 Development environment

In this particular study, for the sake of simplicity we used the rheumatology environment as a development environment. Since it was already granted access to the clinical language model, and since the annotated datasets underwent a time-consuming de-identification to enable their transfer between environments (see above), we could faithfully emulate the proposed workflow presented in Figure 1 while complying to the current rules of the CDW. More generally, we can advocate for the use of such a development environment for future training of machine learning models, together with the trained model being made available through a secure model hub. Access to this environment could be granted only to a limited number of machine learning engineers to limit privacy exposure.

#### 4 Appendix D: Computation of the CCI

Beyond the analysis of each condition, we moreover compared three approaches to compute the CCI: i) using *Quan et al.* formula to aggregate the conditions detected by the **NLP-ML-CLINICAL** pipeline ($CCI_{ML}$), ii) using the same formula to aggregate the conditions detected by the **CLAIM** pipeline ($CCI_{Claim}$) or iii) using direct detection of the CCI score computed by clinicians at the point of care and reported in the clinical notes ($CCI_{Reported}$).[5] For this evaluation we considered an additional sub-cohort composed of the three cohorts' inpatient stays with an admission in an intensive care unit (ICU) department and for which a simple rule-based NLP algorithm detected a CCI reported by the clinicians. The coherence between these scores was assessed by computing $\Delta(CCI)$, the differences of $CCI_{ML}$ with $CCI_{Claim}$ and $CCI_{Reported}$, respectively. Additionally, correlation between CCI computed by two distinct methods were assessed via Spearman correlation coefficient.

The ICU cohort consisted of 46, 1288 and 279 stays for the cardiology, oncology and rheumatology studies respectively. Figure S5 shows the distribution of $\Delta(CCI)$ comparing $CCI_{ML}$ with either $CCI_{Reported}$ or $CCI_{Claim}$. Overall, the distributions have a median (and interquartile range) of 1 (4) and 0 (2), respectively. Specifically, $CCI_{ML}$ and $CCI_{Claim}$ fully agree for 33% of the stays versus 16% for $CCI_{ML}$ and $CCI_{Reported}$. When disagreeing, $CCI_{ML}$ is higher than $CCI_{Claim}$ for 70% of the stays. More generally, CCI computed via two distinct methods show poor correlation, with correlation coefficient of 0.22, 0.41 and 0.38 for $CCI_{ML}$ vs $CCI_{Reported}$, $CCI_{ML}$ vs $CCI_{Claim}$ and $CCI_{Claim}$ vs $CCI_{Reported}$, respectively.

The agreement with $CCI_{ML}$ is still higher for $CCI_{Claim}$ than for $CCI_{Reported}$, having both a smaller interquartile range and a median closer to zero. Most stays featuring a disagreement between $CCI_{ML}$ and $CCI_{Claim}$ have a higher $CCI_{ML}$. This discrepancy could have partially occurred because we added some conditions initially absent to the CCI, such as arterial hypertension in peripheral vascular disease or atrial fibrillation in congestive heart failure. Those conditions are known to be prone to under-coding, thus biasing $CCI_{Claim}$ towards lower values without impacting $CCI_{ML}$ as much. More generally, pairwise correlation is poor regardless of method, with each method exhibiting limitations: $CCI_{ML}$ was computed on patients specifically visiting ICU departments while being built and trained on a more general corpus, which could impair performances. As mentioned above, $CCI_{Claim}$ suffers from coding biases. $CCI_{Reported}$ was computed while patients were in ICU, in an emergency situation where information about each condition was not necessarily available at the time of calculation. Additionally, we lacked information regarding the method and clinical rationale used by the many clinicians who computed these $CCI_{Reported}$. In the end, no method seems to guarantee a ground truth regarding the CCI, prompting one to be cautious about the use of those calculated scores.

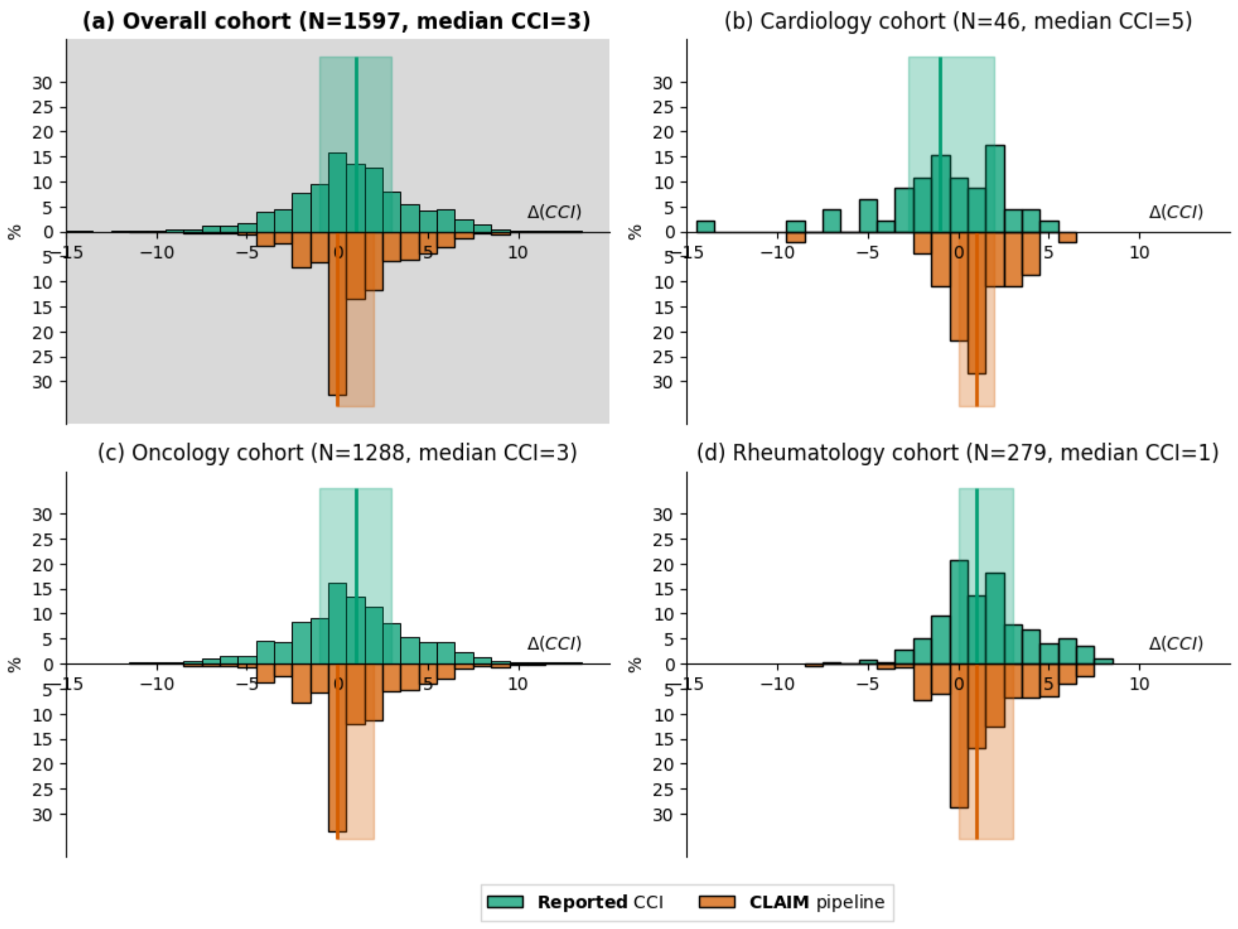


Figure S5: Difference between the Charlson Comorbidity Index obtained using the main **NLP-ML-CLINICAL** pipeline and its value computed either by the clinician at the point of care (green, number of inpatient stays and median score in brackets) or a posteriori from the alternative **CLAIM** pipeline (orange). Distributions, medians and interquartile ranges are indicated by histograms, vertical lines and shaded areas, respectively. The cardiology (b), oncology (c), rheumatology (d) or overall (a) cohorts are considered separately.

#### 5 Appendix E: Additional results

| | Oncology cohort | | Cardiology cohort | | Rheumatology cohort | | Overall cohort | |
|---|---|---|---|---|---|---|---|---|
| **Stay type** | **inpatient** | **outpatient** | **inpatient** | **outpatient** | **inpatient** | **outpatient** | **inpatient** | **outpatient** |
| **Age at admission** | 84.6 (7.3) | 80.9 (6.9) | 61.4 (18.7) | 61.9 (15.2) | 57.5 (19.4) | 53.4 (19.2) | 67.8 (20.0) | 65.4 (18.6) |
| **Gender distribution (M-F)** | 48 - 52 | 48 - 52 | 60 - 40 | 46 - 54 | 50 - 50 | 36 - 64 | 52 - 47 | 43 - 56 |
| **Total annotated records** | 50 | 50 | 50 | 50 | 50 | 50 | 150 | 150 |
| **Myocardial infarction** | 15 (3.5) | 10 (2.1) | 10 (2.7) | 3 (1.3) | 10 (2.7) | 4 (1.5) | 35 (3.1) | 17 (1.8) |
| **Congestive heart failure** | 41 (6.3) | 22 (3.0) | 15 (4.1) | 1 (4.0) | 32 (3.3) | 2 (1.0) | 88 (4.8) | 25 (2.8) |
| **Peripheral vascular disease** | 41 (2.3) | 17 (2.0) | 29 (2.6) | 7 (1.4) | 37 (2.9) | 12 (2.7) | 107 (2.6) | 36 (2.1) |
| **Cerebrovascular disease** | 12 (1.8) | 4 (1.8) | 5 (2.4) | 4 (1.2) | 16 (5.7) | 5 (1.6) | 33 (3.8) | 13 (1.5) |
| **Dementia** | 15 (1.7) | 2 (2.5) | 4 (1.8) | 0 | 9 (3.1) | 3 (1.3) | 28 (2.2) | 5 (1.8) |
| **Chronic pulmonary disease** | 22 (4.2) | 15 (1.7) | 6 (3.5) | 6 (1.5) | 16 (3.4) | 11 (1.5) | 44 (3.8) | 32 (1.6) |
| **Rheumatologic disease** | 1 (1.0) | 3 (1.3) | 3 (2.7) | 2 (1.5) | 19 (4.2) | 15 (2.1) | 23 (3.9) | 20 (2.0) |
| **Peptic ulcer disease** | 7 (1.4) | 0 | 3 (1.7) | 1 (1.0) | 3 (1.3) | 0 | 13 (1.5) | 1 (1.0) |
| **Liver disease** | 0 | 1 (3.0) | 2 (8.0) | 1 (2.0) | 8 (6.5) | 3 (2.3) | 10 (6.8) | 5 (2.4) |
| **Diabetes** | 16 (2.2) | 13 (1.4) | 14 (2.5) | 4 (1.8) | 15 (3.0) | 8 (1.5) | 45 (2.6) | 25 (1.5) |
| **Hemiplegia**$^\star$ | 1 (2.0) | 1 (2.0) | 1 + *10* (6.0) | 0 + *14* (-) | 2 + *15* (1.0) | 0 + *15* (-) | 4 + *25* (2.5) | 1 + *29* (2.0) |
| **Renal disease** | 10 (1.8) | 4 (2.0) | 5 (1.2) | 2 (2.0) | 1 (4.0) | 0 | 16 (1.8) | 6 (2.0) |
| **Solid tumor** | 11 (2.6) | 6 (2.2) | 27 (3.4) | 23 (2.6) | 11 (1.5) | 10 (1.4) | 49 (2.8) | 39 (2.2) |
| **Leukemia** | 2 (3.0) | 1 (4.0) | 3 (1.3) | 3 (2.7) | 4 (3.5) | 3 (1.7) | 9 (2.7) | 7 (2.4) |
| **Lymphoma** | 7 (4.6) | 8 (1.4) | 5 (4.6) | 5 (2.0) | 7 (1.9) | 1 (1.0) | 19 (3.6) | 14 (1.6) |
| **AIDS**$^\star$ | 1 (1.0) | 0 | 1 + *8* (2.0) | 0 + *8* (-) | 0 + *13* (-) | 0 + *9* (-) | 2 + *21* (1.5) | 0 + *17* (-) |
| **Alcohol Consumption** | 14 (1.1) | 1 (1.0) | 13 (1.6) | 1 (2.0) | 18 (1.9) | 3 (1.0) | 45 (1.6) | 5 (1.2) |
| **Tobacco consumption** | 24 (1.5) | 8 (1.4) | 20 (1.6) | 11 (1.1) | 24 (1.6) | 6 (1.3) | 68 (1.5) | 25 (1.2) |
| **Total** | 49 (14.9) | 41 (5.6) | 46 (9.6) | 38 (3.7) | 48 (14.9) | 40 (3.8) | 143 (13.2) | 119 (4.4) |

$^\star$ For those conditions, the number of upsampled notes is also shown *in italics*

Table S4: Composition of the validation dataset sampled in the three disease-specific cohorts without considering qualification variables (i.e., including negated and hypothetical entities, or entities not related to the patient). For each condition, the number of clinical notes with at least one validated entity is shown along with, for those notes, the mean number of validated entities per note (in brackets). For instance, in the cardiology cohort, 15 discharge summaries mention a myocardial infarction and on average 3.5 validated entities are present within each one of them.

| | | F1 | Positive predictive value | Sensitivity |
|---|---|---|---|---|
| **Myocardial infarction** | | 92,3 | 95,3 | 89,5 |
| **Congestive heart failure** | | 92,3 | 91,9 | 92,6 |
| **Peripheral vascular disease** | | 89,3 | 91,6 | 87,1 |
| **Cerebrovascular disease** | | 87 | 87,9 | 86 |
| **Dementia** | | 90,4 | 92,9 | 88,1 |
| **Chronic pulmonary disease** | | 90,4 | 87,9 | 93 |
| **Rheumatologic disease** | | 84,7 | 94,7 | 76,6 |
| **Peptic ulcer disease** | | 100 | 100 | 100 |
| **Liver** | *Mild* | 94,1 | 100 | 88,9 |
| | *Moderate to severe* | 88 | 84,6 | 91,7 |
| | *Any* | 89,9 | 89,1 | 90,7 |
| **Diabetes** | *Without complications* | 89 | 94,2 | 84,4 |
| | *With complications* | 83,3 | 87 | 80 |
| | *Any* | 92,2 | 97,2 | 87,6 |
| **Hemiplegia**⋆ | | - | 91,1 | - |
| **Renal disease** | | 80,6 | 82,9 | 78,4 |
| **Solid tumor** | *Localized* | 91,1 | 93,9 | 88,5 |
| | *Metastatic* | 90,1 | 91,4 | 88,9 |
| | *Any* | 90,9 | 93,3 | 88,6 |
| **Leukemia** | | 94,1 | 97 | 91,4 |
| **Lymphoma** | | 91,4 | 91,4 | 91,4 |
| **AIDS**⋆ | | - | 57,5 | - |
| **Alcohol consumption** | *Present* | 84,7 | 78,1 | 92,6 |
| | *Stopped* | 63,2 | 100 | 46,2 |
| | *Any* | 94,9 | 97,4 | 92,5 |
| **Tobacco consumption** | *Present* | 87,6 | 90,7 | 84,8 |
| | *Stopped* | 94,9 | 94,9 | 94,9 |
| | *Any* | 94,6 | 96,3 | 92,9 |
| **Total** | **Micro average** | 90,6 | 92,3 | 89 |
| | **Macro average** | 90,9 | 92,9 | 89,2 |
| | **Weighted average** | 90,7 | 92,4 | 89,3 |

⋆ For those conditions, only the positive predictive value was computed.

Table S5: Entity-level performances (F1-score, positive predictive value and sensitivity) of the main **NLP-ML-CLINICAL** pipeline used to predict 18 conditions from clinical notes. We considered jointly inpatient and outpatient of the three disease-specific cohorts.

| | | Cardiology cohort | | | | Oncology cohort | | | | Rheumatology cohort | | | |
|---|---|---|---|---|---|---|---|---|---|---|---|---|---|
| | | **F1** | **PPV** | **Sensitivity** | **Specificity** | **F1** | **PPV** | **Sensitivity** | **Specificity** | **F1** | **PPV** | **Sensitivity** | **Specificity** |
| **Myocardial infarction** | | 95,2 | 95,2 | 95,2 | 98,7 | 85,7 | 81,8 | 90 | 97,8 | 94,7 | 100 | 90 | 100 |
| **Congestive heart failure** | | 95,4 | 96,3 | 94,5 | 95,6 | 100 | 100 | 100 | 100 | 96,8 | 93,8 | 100 | 98,8 |
| **Peripheral vascular disease** | | 97,2 | 96,4 | 98,1 | 95,7 | 98,5 | 97 | 100 | 98,5 | 97,3 | 100 | 94,7 | 100 |
| **Cerebrovascular disease** | | 100 | 100 | 100 | 100 | 90,9 | 83,3 | 100 | 98,9 | 90,3 | 87,5 | 93,3 | 97,6 |
| **Dementia** | | 93,3 | 93,3 | 93,3 | 98,8 | 85,7 | 75 | 100 | 99 | 96 | 92,3 | 100 | 98,9 |
| **Chronic pulmonary disease** | | 95,4 | 93,9 | 96,9 | 97,1 | 84,2 | 80 | 88,9 | 97,8 | 97,1 | 94,4 | 100 | 98,8 |
| **Rheumatologic disease** | | 66,7 | 100 | 50 | 100 | 66,7 | 100 | 50 | 100 | 98 | 96,2 | 100 | 98,7 |
| **Peptic ulcer disease** | | 100 | 100 | 100 | 100 | 100 | 100 | 100 | 100 | 100 | 100 | 100 | 100 |
| **Liver** | *Mild* | - | - | - | - | 100 | 100 | 100 | 100 | 100 | 100 | 100 | 100 |
| | *Moderate to severe* | 100 | 100 | 100 | 100 | 100 | 100 | 100 | 100 | 100 | 100 | 100 | 100 |
| | *Any* | 100 | 100 | 100 | 100 | 100 | 100 | 100 | 100 | 100 | 100 | 100 | 100 |
| **Diabetes** | *Without complications* | 97,6 | 100 | 95,2 | 100 | 100 | 100 | 100 | 100 | 90,9 | 100 | 83,3 | 100 |
| | *With complications* | 100 | 100 | 100 | 100 | 100 | 100 | 100 | 100 | 75 | 60 | 100 | 97,9 |
| | *Any* | 98,2 | 100 | 96,4 | 100 | 100 | 100 | 100 | 100 | 100 | 100 | 100 | 100 |
| **Hemiplegia**⋆ | | - | - | - | - | - | 79,3 | - | - | - | 100 | - | - |
| **Renal disease** | | 87 | 100 | 76,9 | 100 | 80 | 66,7 | 100 | 96,8 | 100 | 100 | 100 | 100 |
| **Solid tumor** | *Localized* | 94,7 | 100 | 90 | 100 | 92,9 | 92,9 | 92,9 | 97,2 | 92,3 | 85,7 | 100 | 97,7 |
| | *Metastatic* | 100 | 100 | 100 | 100 | 96,3 | 100 | 92,9 | 100 | 100 | 100 | 100 | 100 |
| | *Any* | 96 | 100 | 92,3 | 100 | 96,4 | 97,6 | 95,2 | 98,3 | 92,9 | 86,7 | 100 | 97,7 |
| **Leukemia** | | 80 | 66,7 | 100 | 99 | 100 | 100 | 100 | 100 | 100 | 100 | 100 | 100 |
| **Lymphoma** | | 100 | 100 | 100 | 100 | 100 | 100 | 100 | 100 | 85,7 | 100 | 75 | 100 |
| **AIDS**⋆ | | - | - | - | - | - | 53,3 | - | - | - | 69,6 | - | - |
| **Alcohol consumption** | *Present* | 100 | 100 | 100 | 100 | 75 | 60 | 100 | 97,9 | 85,7 | 75 | 100 | 99 |
| | *Stopped* | 100 | 100 | 100 | 100 | 50 | 100 | 33,3 | 100 | 85,7 | 100 | 75 | 100 |
| | *Any* | 100 | 100 | 100 | 100 | 100 | 100 | 100 | 100 | 100 | 100 | 100 | 100 |
| **Tobacco consumption** | *Present* | 83,3 | 83,3 | 83,3 | 98,9 | 71,4 | 62,5 | 83,3 | 96,8 | 80 | 85,7 | 75 | 98,9 |
| | *Stopped* | 100 | 100 | 100 | 100 | 95,2 | 100 | 90,9 | 100 | 85,7 | 81,8 | 90 | 97,8 |
| | *Any* | 94,4 | 94,4 | 94,4 | 98,8 | 91,4 | 88,9 | 94,1 | 97,6 | 100 | 100 | 100 | 100 |
| **Total** | **Micro average** | 95,6 | 96,5 | 94,8 | 99,2 | 94,6 | 93 | 96,1 | 99,1 | 96,9 | 96,2 | 97,6 | 99,4 |
| | **Macro average** | 93,7 | 96 | 93 | 99 | 92,5 | 91,9 | 94,9 | 99 | 96,8 | 96,9 | 97,1 | 99,4 |
| | **Weighted average** | 95,5 | 96,7 | 94,8 | 97,8 | 94,6 | 93,9 | 96,1 | 98,7 | 96,9 | 96,4 | 97,6 | 99,3 |

⋆ For those conditions, only the positive predictive value was computed.

Table S8: Stay-level performance of the main **NLP-ML-CLINICAL** stratified by cohort.

| | | F1 | | | Positive predictive value | | | Sensitivity | | |
|---|---|---|---|---|---|---|---|---|---|---|
| | | **NLP-ML-CLINICAL** | **NLP-ML-GENERIC** | **NLP-RB** | **NLP-ML-CLINICAL** | **NLP-ML-GENERIC** | **NLP-RB** | **NLP-ML-CLINICAL** | **NLP-ML-GENERIC** | **NLP-RB** |
| **Myocardial infarction** | | **96,9** | 96,5 | 95,7 | **98,2** | 96,5 | 94,1 | 95,6 | 96,5 | **97,4** |
| **Congestive heart failure** | | **97,5** | 97 | 95,9 | 96,2 | 96,4 | **96,5** | **98,9** | 97,6 | 95,3 |
| **Peripheral vascular disease** | | **95,8** | 95 | 92,8 | **97,6** | 96,7 | 97 | **94,1** | 93,3 | 89 |
| **Cerebrovascular disease** | | 93,8 | 93,6 | **94,6** | 90,9 | 92,6 | **95,6** | **96,8** | 94,6 | 93,5 |
| **Dementia** | | **96,7** | 92,6 | 88,9 | **93,7** | 90,3 | 89,7 | **100** | 94,9 | 88,1 |
| **Chronic pulmonary disease** | | 95,2 | **96,5** | 94,6 | 91,9 | **96** | 93,2 | **98,8** | 97,1 | 95,9 |
| **Rheumatologic disease** | | **95,2** | 92,8 | 93,8 | 95,7 | **96,6** | 91 | 94,7 | 89,4 | **96,8** |
| **Peptic ulcer disease** | | **100** | **100** | 90,9 | **100** | **100** | 88,2 | **100** | **100** | 93,8 |
| **Liver** | *Mild* | **97,1** | **97,1** | 91,4 | **100** | **100** | 94,1 | **94,4** | **94,4** | 88,9 |
| | *Moderate to severe* | **90,9** | 89,2 | 84,6 | 85,4 | **86,8** | 78,6 | **97,2** | 91,7 | 91,7 |
| | *Any* | **92,9** | 91,7 | 86,7 | 89,7 | **90,9** | 83,1 | **96,3** | 92,6 | 90,7 |
| **Diabetes** | *Without complications* | **92,3** | 90,5 | 86,9 | **97,7** | 97,6 | 87,4 | **87,5** | 84,4 | 86,5 |
| | *With complications* | **96,2** | 90,9 | 85,2 | **92,6** | 83,3 | 79,3 | **100** | **100** | [illegible] |
| | *Any* | **93,2** | 90,6 | 86,5 | **96,5** | 93,8 | 85,5 | **90,1** | 87,6 | [illegible] |
| **Hemiplegia*** | | - | - | - | **97,4** | 93,6 | 94,9 | - | - | - |
| **Renal disease** | | **100** | 98,7 | 95,9 | **100** | 97,4 | 97,2 | **100** | **100** | 94,6 |
| **Solid tumor** | *Localized* | **95,6** | 94 | 92,1 | **93,7** | 91,5 | 89,2 | **97,5** | 96,7 | [illegible] |
| | *Metastatic* | 90,9 | **93,3** | 92,3 | 85,4 | **89,7** | 85,7 | 97,2 | 97,2 | **100** |
| | *Any* | **94,5** | 93,9 | 92,1 | **91,7** | 91,1 | 88,4 | **97,5** | 96,8 | 96,2 |
| **Leukemia** | | 97,1 | **100** | 98,6 | **100** | **100** | **100** | 94,3 | **100** | 97,1 |
| **Lymphoma** | | 93,1 | **93,4** | 84 | **93,1** | 89,1 | 75,3 | 93,1 | **98,3** | 94,8 |
| **AIDS*** | | - | - | - | 94 | **95,7** | 92,5 | - | - | - |
| **Alcohol consumption** | *Present* | **94,3** | 86,3 | 83,6 | **96,2** | 91,7 | 82,1 | **92,6** | 81,5 | 85,2 |
| | *Stopped* | **100** | 96 | 91,7 | **100** | **100** | **100** | **100** | 92,3 | 84,6 |
| | *Any* | **96,2** | 89,5 | 86,1 | **97,4** | 94,4 | 87,2 | **95** | 85 | 85 |
| **Tobacco consumption** | *Present* | **92,3** | 85,4 | 76,4 | **93,3** | 88,4 | 65,6 | **91,3** | 82,6 | **91,3** |
| | *Stopped* | **98,7** | **98,7** | 94,6 | **100** | **100** | **100** | **97,4** | **97,4** | 89,7 |
| | *Any* | **95,2** | 91,6 | 83,7 | **96,4** | 93,8 | 77,8 | **94,1** | 89,4 | 90,6 |
| **Total** | **Micro average** | **95,7** | 94,8 | 92,4 | **95,1** | 94,9 | 91,5 | **96,3** | 94,7 | 93,3 |
| | **Macro average** | **95,8** | 94,6 | 91,3 | **95,5** | 94,7 | 90 | **96,2** | 94,6 | 92,9 |
| | **Weighted average** | **95,7** | 94,8 | 92,5 | **95,2** | 94,9 | 91,9 | **96,3** | 94,7 | 93,3 |

* For those conditions, only the positive predictive value was computed.

Table S6: Stay-level performance of the qualification algorithms only of the main **NLP-ML-CLINICAL** pipeline, the **NLP-ML-GENERIC** pipeline and the **NLP-RB** pipeline: algorithms ran on validated entities only.

| Stay type | | F1 | | Positive predictive value | | Sensitivity | | Specificity | |
|---|---|---|---|---|---|---|---|---|---|
| | | inpatient | outpatient | inpatient | outpatient | inpatient | outpatient | inpatient | outpatient |
| **Myocardial infarction** | | 94.9 (89.2 - 98.4) | 87.0 (76.0 - 96.0) | 90.3 (80.6 - 96.8) | 100.0 ( - ) | 100.0 ( - ) | 76.9 (61.3 - 92.3) | 97.5 (95.2 - 99.2) | 100.0 ( - ) |
| **Congestive heart failure** | | 95.7 (91.1 - 98.3) | 97.9 (95.6 - 98.0) | 94.9 (89.7 - 98.4) | 100.0 ( - ) | 96.6 (91.0 - 98.3) | 95.8 (91.6 - 96.0) | 96.7 (93.7 - 98.9) | 100.0 ( - ) |
| **Peripheral vascular disease** | | 97.2 (95.4 - 98.4) | 98.6 (97.1 - 98.6) | 97.8 (95.6 - 98.9) | 97.1 (94.3 - 97.2) | 96.7 (93.3 - 98.9) | 100.0 ( - ) | 96.7 (93.5 - 98.3) | 99.1 (98.3 - 99.1) |
| **Cerebrovascular disease** | | 96.0 (91.6 - 98.0) | 88.9 (75.0 - 90.9) | 92.3 (84.5 - 96.2) | 88.9 (70.0 - 90.9) | 100.0 ( - ) | 88.9 (66.7 - 90.9) | 98.4 (96.9 - 99.2) | 99.3 (97.9 - 99.3) |
| **Dementia** | | 92.3 (85.7 - 94.5) | 100.0 ( - ) | 88.9 (78.6 - 92.9) | 100.0 ( - ) | 96.0 (88.4 - 96.6) | 100.0 ( - ) | 97.6 (95.3 - 98.4) | 100.0 ( - ) |
| **Chronic pulmonary disease** | | 92.8 (86.2 - 97.3) | 96.0 (89.8 - 96.3) | 88.9 (80.5 - 97.3) | 96.0 (87.4 - 96.3) | 97.0 (90.9 - 97.3) | 96.0 (91.2 - 96.3) | 96.6 (94.2 - 99.1) | 99.2 (97.6 - 99.2) |
| **Rheumatologic disease** | | 87.5 (71.4 - 94.4) | 96.8 (89.7 - 97.0) | 93.3 (73.3 - 94.5) | 100.0 ( - ) | 82.4 (62.4 - 94.4) | 93.8 (81.2 - 94.1) | 99.2 (97.1 - 99.2) | 100.0 ( - ) |
| **Peptic ulcer disease** | | 100.0 ( - ) | 100.0 ( - ) | 100.0 ( - ) | 100.0 ( - ) | 100.0 ( - ) | 100.0 ( - ) | 100.0 ( - ) | 100.0 ( - ) |
| **Liver** | *Mild* | 100.0 ( - ) | 100.0 ( - ) | 100.0 ( - ) | 100.0 ( - ) | 100.0 ( - ) | 100.0 ( - ) | 100.0 ( - ) | 100.0 ( - ) |
| | *Moderate to severe* | 100.0 ( - ) | 100.0 ( - ) | 100.0 ( - ) | 100.0 ( - ) | 100.0 ( - ) | 100.0 ( - ) | 100.0 ( - ) | 100.0 ( - ) |
| | *Any* | 100.0 ( - ) | 100.0 ( - ) | 100.0 ( - ) | 100.0 ( - ) | 100.0 ( - ) | 100.0 ( - ) | 100.0 ( - ) | 100.0 ( - ) |
| **Diabetes** | *Without complications* | 96.4 (92.5 - 98.2) | 96.8 (89.5 - 97.0) | 100.0 ( - ) | 100.0 ( - ) | 93.1 (86.1 - 96.6) | 93.8 (80.9 - 94.1) | 100.0 ( - ) | 100.0 ( - ) |
| | *With complications* | 94.1 (87.3 - 94.7) | 92.3 (83.1 - 93.3) | 88.9 (77.5 - 90.0) | 85.7 (71.1 - 87.5) | 100.0 ( - ) | 100.0 ( - ) | 99.3 (98.6 - 99.3) | 99.3 (98.6 - 99.3) |
| | *Any* | 98.6 (95.7 - 98.7) | 100.0 ( - ) | 100.0 ( - ) | 100.0 ( - ) | 97.3 (91.8 - 97.4) | 100.0 ( - ) | 100.0 ( - ) | 100.0 ( - ) |
| **Hemiplegia*** | | - | - | 82.8 (71.4 - 93.4) | 96.3 (88.8 - 96.4) | - | - | - | - |
| **Renal disease** | | 85.7 (72.0 - 93.8) | 83.3 (44.4 - 92.3) | 92.3 (81.7 - 93.8) | 71.4 (28.6 - 85.7) | 80.0 (60.0 - 93.8) | 100.0 ( - ) | 99.3 (98.5 - 99.3) | 98.6 (96.6 - 99.3) |
| **Solid tumor** | *Localized* | 96.0 (89.4 - 98.0) | 90.2 (83.2 - 94.7) | 92.3 (80.8 - 96.2) | 92.0 (80.7 - 96.2) | 100.0 ( - ) | 88.5 (80.0 - 96.4) | 98.4 (96.1 - 99.2) | 98.4 (96.1 - 99.2) |
| | *Metastatic* | 100.0 ( - ) | 92.3 (72.0 - 93.3) | 100.0 ( - ) | 100.0 ( - ) | 100.0 ( - ) | 85.7 (56.4 - 87.5) | 100.0 ( - ) | 100.0 ( - ) |
| | *Any* | 97.2 (94.2 - 98.6) | 93.8 (86.6 - 97.1) | 94.6 (89.1 - 97.3) | 96.8 (89.9 - 97.1) | 100.0 ( - ) | 90.9 (81.2 - 97.1) | 98.3 (96.6 - 99.1) | 99.1 (97.5 - 99.1) |
| **Leukemia** | | 94.1 (71.4 - 94.7) | 100.0 ( - ) | 88.9 (55.6 - 90.0) | 100.0 ( - ) | 100.0 ( - ) | 100.0 ( - ) | 99.3 (97.2 - 99.3) | 100.0 ( - ) |
| **Lymphoma** | | 95.7 (85.7 - 96.0) | 100.0 ( - ) | 100.0 ( - ) | 100.0 ( - ) | 91.7 (75.0 - 92.3) | 100.0 ( - ) | 100.0 ( - ) | 100.0 ( - ) |
| **AIDS** | | - | - | 55.6 (44.2 - 70.4) | 65.4 (49.9 - 80.8) | - | - | - | - |
| **Alcohol consumption** | *Present* | 76.9 (40.0 - 93.3) | 100.0 ( - ) | 62.5 (25.0 - 87.5) | 100.0 ( - ) | 100.0 ( - ) | 100.0 ( - ) | 97.9 (95.9 - 99.3) | 100.0 ( - ) |
| | *Stopped* | 80.0 (50.0 - 94.1) | - | 100.0 ( - ) | - | 66.7 (33.3 - 88.9) | - | 100.0 ( - ) | - |
| | *Any* | 100.0 ( - ) | 100.0 ( - ) | 100.0 ( - ) | 100.0 ( - ) | 100.0 ( - ) | 100.0 ( - ) | 100.0 ( - ) | 100.0 ( - ) |
| **Tobacco consumption** | *Present* | 66.7 (44.2 - 88.1) | 90.0 (70.4 - 95.2) | 70.0 (49.8 - 90.9) | 81.8 (54.3 - 90.9) | 63.6 (36.1 - 91.7) | 100.0 ( - ) | 97.8 (96.4 - 99.3) | 98.6 (96.5 - 99.3) |
| | *Stopped* | 93.9 (84.3 - 96.2) | 94.1 (87.3 - 94.7) | 92.0 (75.9 - 96.2) | 100.0 ( - ) | 95.8 (89.4 - 96.2) | 88.9 (77.5 - 90.0) | 98.4 (95.4 - 99.2) | 100.0 ( - ) |
| | *Any* | 94.3 (89.5 - 95.8) | 97.3 (88.2 - 97.4) | 94.3 (88.8 - 97.1) | 94.7 (78.9 - 95.0) | 94.3 (85.6 - 97.1) | 100.0 ( - ) | 98.3 (96.6 - 99.1) | 99.2 (97.0 - 99.2) |
| **Total** | **Micro average** | 95.5 (93.8 - 95.9) | 96.2 (93.9 - 96.9) | 94.8 (92.8 - 95.6) | 96.9 (93.7 - 97.4) | 96.2 (94.2 - 96.9) | 95.6 (93.0 - 96.6) | 98.8 (98.3 - 99.0) | 99.7 (99.4 - 99.7) |
| | **Macro average** | 95.1 (92.7 - 95.7) | 96.2 (93.0 - 97.0) | 94.8 (92.2 - 95.3) | 96.6 (93.2 - 97.5) | 95.7 (93.2 - 96.4) | 96.4 (94.2 - 97.3) | 98.6 (98.1 - 98.9) | 99.7 (99.3 - 99.7) |
| | **Weighted average** | 95.5 (93.7 - 95.9) | 96.2 (94.0 - 96.9) | 94.9 (93.1 - 95.7) | 97.2 (94.9 - 97.5) | 96.2 (94.2 - 96.9) | 95.6 (93.0 - 96.6) | 98.0 (97.2 - 98.5) | 99.5 (99.2 - 99.5) |

In case of metrics at 100, no bootstraping was performed.

* For those conditions, only the positive predictive value was computed.

Table S7: Stay-level performances (F1-score, positive predictive value, sensitivity and specificity) of the main **NLP-ML-CLINICAL** pipeline used to predict 18 conditions from clinical notes stratified by stay type. 95% confidence intervals were estimated by bootstrapping.

### 6 Appendix F: Annotation guideline

#### 6.1 Annotation content

Each entity should be annotated along with, when relevant, its severity, the start and end of the entity in the text, and qualifiers deduced from its context (negation, family, hypothesis). Below are a few examples with their corresponding annotation in Table S9.

**Examples:**

- M. DUPONT est **diabétique**. (*Mr DUPONT is **diabetic**.*)
- Probable **SAPL** chez le père du patient. (*Probable **APS** in the patient's father.*)
- A été traité à **AVC**[1] en 2020. (*Was treated at **AVC** hospital in 2020*)

| | **Label** | **Status** | **Negation** | **Family** | **Hypothesis** | **Start** | **End** | **Anon. Text** |
|---|---|---|---|---|---|---|---|---|
| **Definition:** | The associated comorbidity | The status of the comorbidity | Is the entity negated ? | Is the entity referring to someone different that the patient | Is the entity uncertain ? | Start position of the entity | End position of the entity | Snippet with identifying traits replaced |
| **Example 1** | Diabetes | Without complications | NO | NO | NO | 15 | 25 | M. MARTIN est diabétique |
| **Example 2** | Peripheral vascular disease | Present | NO | YES | YES | 9 | 13 | Probable SAPL chez le père du patient |
| **Example 3** | Cerebrovascular disease | Absent | NO | NO | NO | 15 | 18 | A été traité à AVC en 2010 |

Table S9: Details of the annotation of a few examples.

#### 6.2 General considerations

**Acute manifestations of diseases are not considered**:

- Le patient est passé aux urgences suite à une insuffisance rénale aigüe (*The patient was admitted to emergency with acute renal failure.*) → **NO**
- Le patient a un insuffisance rénale de stade 4 (*Patient has stage 4 renal failure*) → **YES**

**Are not labeled terms specific to the semiology of radiology and medical imaging, nor the terms present in the physical examination**. Indeed, the scope of the project is only to focus on the certain diagnostic already made and explicitly mentioned in the text, not to predict one:

- IRM cérébrale: présence d'une petite lacune lenticulaire (*Brain MRI: presence of a small lenticular lacuna*) → **Not labeled as cerebrovascular disease.**

**Annotation should be performed at the entity level, i.e. by considering its local context only**:

[1] AVC is an acronym for stroke, but also the acronym of one of AP-HP's hospitals.

For instance, let us consider the following snippet:

- Le patient est suivi pour un diabète (1) de type II. Il a également [...] et une rétinopathie diabétique (2). (*The patient is being followed for type II diabetes (1). He also has [...] and diabetic retinopathy (2)*)

In this case, the status of extraction (1) should be set to *without complications* (nothing, in the immediate vicinity of the entity, mentions a complication), while the status of extraction (2) should be *with complications*.

**In the case of comorbidities with non-binary status, if the status isn't explicitly mentioned, it should be set to its default value:**

- Le patient est atteint d'un cancer du poumon (*Patient has lung cancer*) → ***Solid tumor* set to *localized***

| Label | Description |
|---|---|
| ADRESSE | Street address, e.g., 33 boulevard de Picpus |
| DATE | Any absolute date other than a birthdate |
| DATE_NAISSANCE | Birthdate |
| HOPITAL | Hospital name, e.g., Hôpital Rothschild |
| IPP | Internal AP-HP identifier for patients, displayed as a number |
| MAIL | Email address |
| NDA | Internal AP-HP identifier for visits, displayed as a number |
| NOM | Any last name (patients, doctors, third parties) |
| PRENOM | Any first name (patients, doctors, etc) |
| SECU | Social security number |
| TEL | Any phone number |
| VILLE | Any city |
| ZIP | Any zip code |

Table S10: Types of identifying entities replaced by the de-identification process

#### REFERENCES


[1] Emmanuelle Kempf, Guillaume Lamé, Richard Layese, et al. New cancer cases at the time of sars-cov2 pandemic and related public health policies: a persistent and concerning decrease long after the end of the national lockdown. *European Journal of Cancer*, 150:260–267, 2021.

[2] Romain Bey, Romain Goussault, François Grolleau, et al. Fold-stratified cross-validation for unbiased and privacy-preserving federated learning. *Journal of the American Medical Informatics Association*, 27(8):1244–1251, 2020.

[3] Wendy W Chapman, Will Bridewell, Paul Hanbury, et al. A simple algorithm for identifying negated findings and diseases in discharge summaries. *Journal of biomedical informatics*, 34(5):301–310, 2001.

[4] Basile Dura, Perceval Wajsburt, Thomas Petit-Jean, et al. EDS-NLP: efficient information extraction from French clinical notes, July 2023.

[5] Hude Quan, Bing Li, Chantal M Couris, et al. Updating and validating the charlson comorbidity index and score for risk adjustment in hospital discharge abstracts using data from 6 countries. *American journal of epidemiology*, 173(6):676–682, 2011.
